# Prediction of ADHD diagnosis using brief, low-cost, clinical measures: a competitive model evaluation

**DOI:** 10.1101/2021.12.23.21268330

**Authors:** Michael A. Mooney, Christopher Neighbor, Sarah Karalunas, Nathan F. Dieckmann, Molly Nikolas, Elizabeth Nousen, Jessica Tipsord, Xubo Song, Joel T. Nigg

## Abstract

Proper diagnosis of ADHD is costly, requiring in-depth evaluation via interview, multi-informant and observational assessment, and scrutiny of possible other conditions. The increasing availability of data may allow the development of machine-learning algorithms capable of accurate diagnostic predictions using low-cost measures. We report on the performance of multiple classification methods used to predict a clinician-consensus ADHD diagnosis. Classification methods ranged from fairly simple (e.g., logistic regression) to more complex (e.g., random forest), and also included a multi-stage Bayesian approach. All methods were evaluated in two large (N>1000), independent cohorts. The multi-stage Bayesian classifier provides an intuitive approach that is consistent with clinical workflows, and is able to predict ADHD diagnosis with high accuracy (>86%)—though not significantly better than other commonly used classifiers, including logistic regression. Results suggest that data from parent and teacher surveys is sufficient for high-confidence classifications in the vast majority of cases using relatively straightforward methods.

## INTRODUCTION

The accurate diagnosis of attention-deficit/hyperactivity disorder (ADHD) is critically important given evidence of both over- and under-treatment (Costello et al. 2014; Simon et al. 2015; Massuti et al. 2021) of this costly condition. A full evaluation of ADHD, requiring significant time and resources, involves a structured or semi-structured clinical interview, standardized ratings from parent and teachers, and evaluation of impairment, as well as comorbid conditions that might better explain the diagnosis (APA 2013; Committee on Quality Improvement, Subcommittee on Attention-Deficit/Hyperactivity Disorder 2000; National Guideline Centre (UK) 2018). Yet, surveys suggest that the majority of providers in primary care faced with evaluating ADHD report insufficient knowledge or resources to carry out a full evaluation (Adler et al. 2009; Faraone et al. 2004) leading to efforts to develop additional resources (Loskutova et al. 2021), and perhaps contributing to concerns about diagnostic accuracy.

Other approaches to assist diagnostic evaluation are also possible and are under development using various contemporary computational tools. For example, computerized adaptive testing is a sophisticated method that uses item response theory and large item banks to develop rapid assessment tools (Gibbons et al. 2016; 2020). A similar approach was used to develop the ongoing NIMH PROMIS scales (Irwin et al. 2010). A potentially simpler machine-learning approach is to create a prediction algorithm that clinicians can adapt to their existing methods, relying on standard clinical tools.

This later approach, which we pursue here, has yet to be strongly evaluated for ADHD. The first and most basic challenge is to differentiate the clinical diagnosis (in this case, ADHD) from cases with no clinical diagnosis. The first question within that domain is whether advanced machine learning classifiers can outperform simple algorithms in accurately identifying ADHD, at sample sizes that may be typically available in local clinics. This paper tackles that challenge, by developing and evaluating multiple machine-learning classifiers, ranging in complexity, to differentiate ADHD from non-ADHD youth in a population with a moderate ADHD base rate. Once this is solved, the next steps are to address differentiation of ADHD from other disorders and prediction of clinical course to guide treatment decisions, while considering different settings’ base rates.

The effort to use machine learning classifiers to identify clinical cases of psychiatric disorders on the basis of low-cost clinical data has been surprisingly sparse. Youngstrom *et al*. (Youngstrom et al. 2018) reported on a set of studies attempting to diagnose cases of bipolar disorder in a competitive modeling environment, similar to our approach here for ADHD. They reported that complex machine learning algorithms, in that case Least Absolute Shrinkage and Selection Operation (LASSO) regression, did not markedly outperform the more familiar logistic regression classifier when validated on independent data. Scrutiny of other mental health conditions along similar lines has been limited. One study of ADHD used a computerized interview and teacher and parent rating scales to establish ADHD diagnosis and then related teacher ratings to establish prediction, but without cross-validation (Öztekin et al. 2021). They found no added benefit of MRI measures.

Two questions are salient. First, can brief measures aided by machine-learning algorithms accurately reproduce clinical diagnoses achieved by time-consuming comprehensive semi-structured clinical interviews, or the even more authoritative clinical best-estimate diagnosis? If so, the potential cost savings would be substantial. Second, if the answer to the first question is yes, do complex contemporary machine learning algorithms outperform simpler and more familiar linear models in such prediction at the sample sizes commonly available to local clinics and research studies? This is a methodological question that we also undertake.

To the relative neglect of simple rating scales, much of the recent literature on the prediction of ADHD or other psychiatric diagnosis has focused on brain imaging measures, with dozens of studies but with generally small samples, weak accuracy, or overfitting concerns (limited cross validation) (Rashid and Calhoun 2020). Indeed, it is now known that effect sizes for brain imaging metrics are small (i.e., they explain only a very small fraction of variance in behavioral measures) (Marek et al. 2020), and the cost of MRI renders it currently sub-optimal for a brief, cost reducing ADHD diagnostic method. Likewise, genetic risk factors (e.g., polygenic risk scores or individual variants) still have little utility in terms of clinical diagnosis/prognosis (Ronald, Bode, and Polderman 2021; Martin, Kanai, et al. 2019; Martin, Daly, et al. 2019; Palk et al. 2019), although they may before long be added to a diagnostic algorithm in some form.

In general, studies using machine learning to predict ADHD diagnosis have been conducted with small samples, have lacked external validation (Bledsoe et al. 2016; Wodka et al. 2008; Dvorsky et al. 2016; Duda et al. 2016; Mueller et al. 2010; Christiansen et al. 2020) and have neglected simpler, lower cost algorithms that might enhance clinical efficiency.

Although the NIMH PROMIS project has developed brief measures (Cella et al. 2007), and computer-assisted assessment is emerging (Hall et al. 2016; Nikolas, Marshall, and Hoelzle 2019; Slobodin, Yahav, and Berger 2020), it remains unclear what an optimal degree of density of assessment would be for ADHD diagnosis in a clinical setting. Conceptually, several considerations also bear mention. Parent and teacher ratings are strongly recommended for ADHD evaluation, yet these often do not agree. A parent interview may yield different responses than a rating scale. Rating scales utilize clinical “cut off scores” for ADHD that may or may not be optimal for individual prediction. Finally, cognitive tests remain controversial as ancillary information for ADHD evaluation, yet it is unknown whether they can aid a diagnostic algorithm secondarily. For example, Nikolas *et al*. (2019) reported that in a simple regression model, certain cognitive measures enhanced ADHD prediction accuracy over and above rating scales (Nikolas, Marshall, and Hoelzle 2019). In particular, we examined the relative merits of (a) parent ratings alone, (b) parent + teacher ratings, and (c) parent + teacher + cognitive testing in predicting both a gold standard best-estimate diagnosis and a clinician structured interview diagnosis of ADHD. Relatedly, it has been unclear whether advanced non-linear modeling (“machine learning”) is superior to traditional logistic regression in creating a prediction score, and whether any will outperform a simple rule-based decision aid (2-level decision tree). Hence, we report here on potential optimal assessment depth, as well as a competitive evaluation of multiple classification methods, in a proof-of-concept paper.

Informal Bayesian logic—weighing new information in the context of existing evidence— is central to clinical decision making (Gill, Sabin, & Schmid 2005). Because of this, we evaluate a multi-stage Bayesian prediction approach (a tree-augmented naïve Bayes classifier) to roughly parallel the sequential decision-making process that a time-and cost-conscious clinician uses. In the case of clinical evaluation for ADHD, the decision making would typically start with a brief low-cost assessment (e.g., a single parent symptom checklist), then proceed to decisions about whether to add additional measures such as obtaining teacher ratings or cognitive testing, and so on. Thus, our sequence starts with the most easily-obtained measures (parent rating scales) and proceeds to teacher ratings, and then laboratory cognitive test measures. In addition to the tree-augmented naïve Bayes classifier, we implemented a competitive modeling logic with the following classifiers, varying in computational complexity: a simple 2-level decision tree, standard logistic regression, regularized logistic regression, support vector machine, unrestricted-depth decision tree, random forest, and gradient boosted decision trees. All classification methods were evaluated in two large (N>1000), well-characterized, case-control cohorts to evaluate robustness and generalizability.

## MATERIALS AND METHODS

### Sample Size and Participants

Methodologically, the relation between a machine learning model and sample size is highly complex, depending on many factors, such as model nonlinearity and complexity, sample noise, dependency among the samples, and effectiveness of model optimization process. It has been theoretically derived and empirically demonstrated (Bishop 2006; Abu-Mostafa, Magdon-Ismail, and Lin 2012; Goodfellow, Bengio, and Courville 2016) that complex machine learning models, such as multi-layer deep learning neural networks, are capable of capturing the complex, nonlinear relations between the model inputs and outputs. However, they require a large amount of data to train, due to the large number of model parameters to specify without overfitting. On the other hand, simple models, such as logistic regression models, can still be the appropriate model for a given data set, in cases of modest sample size, highly correlated samples (which reduces the effective sample size), noisy samples, and close-to-linear input-output relations. Thus, evaluation is necessary for the particular question at hand. Here, we opted for well-characterized samples that had sufficient sample size to test the question, that compared well to other similar efforts in the literature, and that might be similar to what most real settings would have available to train a local classifier.

The Oregon-ADHD-1000 is a community recruited case-control cohort (N=1423) of youth age 7-11 years (Karalunas et al. 2017; Nigg et al. 2018; 2020; Mooney et al. 2021; 2020). ADHD was deliberately oversampled to ensure an adequate range of clinical variation and of actual clinical cases in the data set. (We consider base rate issues later). To preserve the representativeness of the sample, we did not oversample for sex or other demographics, and these were not included in our algorithm to mimic clinician context. Human subjects and ethics approval was obtained from the local University Institutional Review Board. A parent/legal guardian provided written informed consent, and children provided written assent.

Recruitment was conducted by community outreach and a case-finding procedure to identify cases in the community, regardless of whether they had sought treatment or been previously diagnosed. After screening, a state-of-the-art, multi-informant, multi-method research diagnostic evaluation was conducted (see below). Children were excluded for disallowed medications (Supplemental Table S1), history of seizures or head injury, psychosis, mania, current major depressive episode, Tourette’s syndrome, autism and IQ<80. An ADHD assignment required all DSM-5 criteria to be met including parent-teacher convergence (both having either above-threshold rating scale scores or symptom counts). To increase the difficulty of the diagnostic case and make it more similar to real world decisions, the sample retained youth who were subthreshold for ADHD (5 symptoms + impairment) and those who had symptoms or impairment explained by other disorders or had situational ADHD symptoms (home or school problems only). As explained below, sensitivity analyses evaluated the effect of these cases.

Replication in a completely independent sample was evaluated with the Michigan-ADHD-1000 (Nikolas and Nigg 2013; 2015). It is a cohort of 1064 youth ages 6-21 years, with the same recruitment and assessment procedures, but in a very different demographic population (central Michigan versus northwest Oregon). Its inclusion enabled a test of generalizability and performance in a completely independent sample to control any conclusions that might derive from single-sample over-fitting.

### ADHD Gold-Standard Diagnosis

In both cohorts, diagnostic assignment followed a standard protocol. It included standardized, nationally-normed rating scales from parent and teacher, parent semi-structured clinical interview by a trained clinician with acceptable inter-clinician reliability and validity, child intellectual testing with carefully trained and supervised psychometrician administrators, and clinical observation and notes from two research assistants. Then, a best-estimate diagnostic team of two experienced clinicians (board certified child psychiatrist and a child clinical psychologist) reviewed all available data and arrived at a best-estimate diagnostic assignment, considering whether symptoms were better explained by an associated condition and whether impairment was severe enough to warrant diagnosis. The two clinicians agreed well (kappa>0.80) on ADHD assignment. Their consensus rating became the gold standard assignment.

These clinician consensus diagnoses, which we refer to as *best-estimate* diagnoses, were used as the ground-truth classifications. The best-estimate team classified each subject as ADHD, control, subthreshold, or other. The *subthreshold* category were children judged by the consensus best-estimate team to have impairing ADHD symptoms but insufficient to meet full diagnostic criteria. The *other* category included subthreshold cases in which only one reporter saw symptoms, or cases with impairment that was judged not due to ADHD symptoms. Non-ADHD cases had 4 or fewer ADHD symptoms and had never been identified or treated for ADHD. Other psychopathology was free to vary in these community volunteers, although we excluded children with mania, a history of probable or definite psychosis, intellectual delay, or autism spectrum disorder or neurological injury.

For our primary analyses we collapse the children into two classes (ADHD, and non-ADHD); the subthreshold and other cases were all labeled as non-ADHD for that primary analysis. In sensitivity analyses we explored the effect of this type of decision versus a broader ADHD definition that included subthreshold cases and also how inclusion or exclusion of these cases from the training set affected classification learning.

The best-estimate team used some of the rating scales that were included in our low-cost classification models, our first analysis focused on whether those results could be approximated with a subset of their information. However, to control the potential “double-dip” of that approach, we also evaluated classifiers trained to predict diagnosis based on the Kiddie Schedule for Affective Disease and Schizophrenia-version E modified for DSM-IV (at the time of data collection) and checked for DSM-5 compliance (after DSM-5 was published). The KSADS-E was administered by a masters-degree or higher clinician (either masters in social work, in counseling, or in clinical psychology) trained to reliability and validity with a master coder who was in turn trained to validity with an outside expert. These interviewers all achieved adequate inter-rater reliability (kappa>0.80) with the master trainer. Recordings of their interviews were regularly reviewed by senior clinicians to prevent administration drift.

### Predictive Measures

The initial modeling used the Oregon cohort. The following symptom measures were used as predictors in the classifiers: parent reported inattention and hyperactivity symptom counts from the *ADHD Rating Scale* (ADHDRS) (DuPaul et al. 1998) parent reported inattention, hyperactivity, executive functioning, learning problems, aggression, and peer relations scales from the *Conners Rating Scale 3*^*rd*^ *edition* (Conners-3) (Conners 2008), and teacher reported inattention and hyperactivity symptom counts from the ADHDRS. All parent and teacher reported measures were converted to T-scores to adjust for age and gender and again to simulate clinical decision making. These symptom measures or others like them are often the first pieces of information used by a clinician when evaluating a child for ADHD.

In addition, cognitive measures from the following laboratory tests were used as predictors: Stop-Go task (Nigg 1999; Schachar et al. 1995); Spatial Span forward and backward (De Luca et al. 2003); Digit Span forward and backward from the Wechsler Intelligence Scale for Children, Fourth Edition (WISC-IV) (Wechsler 2003); Delis, Kaplan, and Kramer (DKEF) (Delis, Kaplan, & Kramer 2001) version of the Stroop task (color, word, and color-word conditions); and DKEF Trail Making test (number sequencing and number-letter sequencing conditions).

For the Michigan cohort, parent reported symptoms were assessed with the ADHDRS (same as the Oregon cohort), and the cognitive problems and hyperactivity-impulsivity scales from the Revised Conners Parent Rating Scale (Conners-R) (Conners et al. 1998), due to the earlier era of data collection prior to publication of the Conners-3. Teacher reported inattention and hyperactivity symptoms were available from the ADHDRS. Again, these symptom measures were converted to T-scores to simulate clinician decision making.

All of the cognitive measures available in the Oregon cohort were also available for the Michigan cohort, although a different version of the Trail Making test was used in Michigan (Reitan and Wolfson 1985)). **Supplemental Table S2** shows all predictive features available in each data set.

### Data Cleaning / Pre-processing

Subjects without a best-estimate team diagnosis and those missing >80% of predictive features were excluded, leaving a total N=1423 and N=1057 subjects for analyses in the Oregon and Michigan cohorts, respectively. KSADS diagnoses were available for 1422 in the Oregon cohort and 1055 in the Michigan cohort.

For the primary analyses in the Oregon cohort, a stratified split of the data was done to maintain equal proportions of ADHD and non-ADHD cases in the sub-samples used for training (75%) and testing (25%) the classifiers. All variables were standardized (mean=0, standard deviation=1) using the StandardScaler method in the Scikit-learn Python package (Pedregosa et al. 2011). The test data was standardized relative to the mean and standard deviation of the training set, to bring the test data into the same range without data leakage from the test set.

### Missing Data Imputation

For machine learning applications, addressing missing data must be done before the optimal predictive model is known (i.e., prior to classifier training). Because of this, model-based methods for handling missing data (e.g., maximum likelihood or regression), which depend on assumptions about the distributions of variables or the relationships between variables, are typically not appropriate. The best approach for machine learning tasks is unknown (and likely data set-specific), but almost certainly listwise deletion is suboptimal. Here, a k-nearest neighbor (KNN) imputation approach, implemented with the KNNImputer method (number of neighbors, k=7) in Scikit-learn, was used to impute missing values. The imputer model was fit with the training data only, to avoid data leakage from the test set, and was then applied to both the training and test sets. KNN imputation has been used in similar applications and has outperformed model-based methods and listwise deletion in terms of classification performance (Jerez et al. 2010). Alternative imputation methods were tested in simulated data (see Supplemental Materials), but provided little advantage at the cost of interpretive difficulty, hence we relied on the KNN approach here.

### Classification Methods

We performed t-Distributed Stochastic Neighbor Embedding (t-SNE) (van der Maaten and Hinton 2008) on the training data set for preliminary data exploration and visualization of how well the predictive features are able to separate the class labels. The TSNE method implemented in Scikit-learn was used with the following parameters: perplexity=30, distance=Euclidean.

#### Bayesian Classifier

As noted earlier, to attempt to simulate the process of clinical decision making so that the results would, in theory, be readily adapted to clinical use, we implemented a Bayesian classifier. Here, we chose a multi-stage tree-augmented naïve Bayes (TAN) classifier. This model was chosen because it models dependencies between predictive features, which we know exist. This is in contrast with standard naïve Bayes classifiers, which assume independence among predictors. The TAN classifiers were implemented using the bnclassify R package (Mihaljević, Bielza, and Larrañaga 2018). This multi-stage approach produces an initial prediction (class probability) based on a small subset of predictors. If a subject’s class probability is below a specified threshold (i.e., low-confidence), additional predictors are added to the classifier and the class probability is updated in subsequent stages. This is illustrated in Figure 1. We conducted a 4-stage approach as follows: Stage 1: parent ADHDRS; stage 2: parent Conners; stage 3: teacher ADHDRS; stage 4: cognitive measures (see Predictive Measures above).

**Figure 1.**
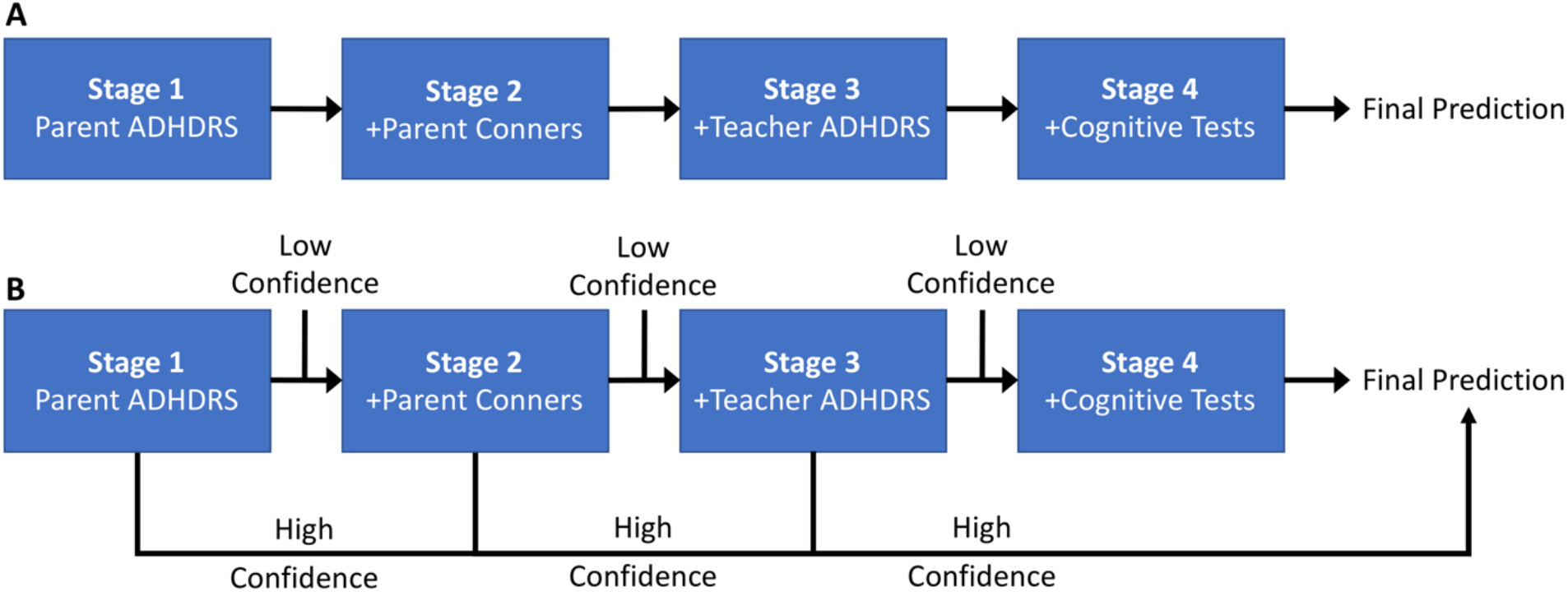
Schematic of the multi-stage classification approach. Each step in the process is a tree-augmented naïve Bayes (TAN) classifier. At each stage, a subject’s prior class probability is updated based on a subset of predictors, and a posterior class probability is produced. In (A), each subject is carried through all stages and the final prediction is based on all data. In (B), if a subject’s posterior probability is above a specified threshold (>0.9; i.e., high-confidence) at any Stage, the final prediction is made at that point and subsequent Stages are skipped.

This multi-stage procedure was evaluated in two ways: (1) the final classification for all participants from stage 4 (i.e., all participants are classified using all predictive features; we refer to this as ‘TAN-stage4’; Figure 1A), or (2) treating an individual’s final classification as the earliest high-confidence classification (i.e., some children are classified using only a subset of predictive features; we refer to this as ‘TAN-earliest’; Figure 1B).

All predictive features were discretized before being input into the TAN classifier—that is, the continuous measures were summarized into a finite set of bins. This was necessary because the TAN classifier depends on the calculation of conditional probability tables from the training data. After standardizing and imputing the data, a z score of +/-1.0 was equivalent to a one-standard deviation position above or below the sample mean, roughly approximating what a clinician might use to gauge whether or not to further pursue a problem (Conners 2003; 2008). In this way, the continuous measures were discretized into four bins: (*x*≤ −1), (−1 > *x*≤ 0), (0 < *x*≤ 1), (*x*> 1). However, alternative methods were evaluated in sensitivity analyses and results, which were generally supportive of the approach chosen, are included in the Supplemental Materials.

#### Competitive Modeling

Using a competitive modeling framework, we compared the performance of multiple classification methods ranging in complexity. We first implemented two relatively simple classification methods, unregularized logistic regression (LR-unregularized) and a decision tree limited to two levels (DT-simple), representing a simple algorithm that could easily be implemented in a clinic unaided by machine learning.

These methods were compared to the multi-stage TAN classifier (described above) and five other advanced methods: regularized logistic regression (LR), unrestricted-depth decision tree (DT), random forest (RF), support vector machine (SVM), and gradient boosted decision trees (GBDT). These classifiers were chosen because many have been used previously in similar applications (Duda et al. 2016; Bledsoe et al. 2016; Dvorsky et al. 2016), they span a reasonable range in terms of complexity and interpretability, and all are easily implemented using the popular Scikit-learn Python package (Pedregosa et al. 2011). Finally, we evaluated an ensemble classifier, which made classifications based on the average class probability across all of the six advanced classifiers (TAN, LR, DT, SVM, RF, and GBDT).

The Bayesian TAN classifier can naturally accommodate a multi-stage approach, by updating the class prior probability at each stage. While other methods could be adapted to a step-wise approach by building independent classifiers with different subsets of features, it is not clear the best way to incorporate information from one step to the next (i.e., the Bayesian logic is not automatic for these other methods). Therefore, we focus on the multi-stage approach for the TAN classifier only. Recognizing this fundamental difference between the methods, we later discuss the relative importance of specific predictive features, and prediction confidence.

#### General procedure for machine learning models

A select set of classifier hyperparameters (Supplemental Table S3) were optimized using a grid search with 5-fold cross-validation as implemented in Scikit-learn (Olson et al. 2018). Default values were used for all other parameters. The hyperparameters that resulted in the highest mean cross-validation accuracy were then used for all subsequent classifier comparisons.

The Oregon cohort data set was split into training and test sets as described above (“Data Cleaning / Pre-processing” section) and the performance of each classifier was evaluated in three ways. First, we performed 5-fold cross validation within the Oregon cohort training set (i.e., mean performance across the 5 folds). Second, we trained the classifiers on the Oregon training set and then classified participants in the hold-out (Oregon) *test set*. Finally, to evaluate external cross-validation and generalizability, we training classifiers on the full Oregon cohort and then classified participants in the independent Michigan cohort. Accuracy, sensitivity (ADHD being the positive class), specificity, positive predictive value (PPV), and area under the receiver operating characteristic curve (AUC-ROC) are reported for all classifiers. PPV was calculated assuming both a 5% prevalence (PPV_5_) of ADHD in the general population (Song, Dieckmann, and Nigg 2019) (as might be seen in a general pediatrics practice) and a 50% prevalence (PPV_50_) (which may be more representative of the population seen in many outpatient psychiatric settings).

Differences in the 5-fold cross-validation accuracy among classifiers was tested with a paired T-test. Given we were primarily interested in determining whether the more advanced methods outperformed unregularized logistic regression, we used a p-value threshold of 0.0071 (a Bonferroni correction for seven comparisons: TAN, LR, DT, DT-simple, SVM, RF, and GBDT) to denote significance. Significant differences observed for the 5-fold cross validation performance were confirmed with an additional 2-fold cross validation repeated 5 times (5×2-fold cross validation) (Dietterich 1998).

## RESULTS

### Description of Samples and cohorts

An overview of the datasets used for training and testing the ADHD classifiers in the Oregon-ADHD-1000 and for the generalizability analysis in the Michigan-ADHD-1000 is provided in Table 1. For primary analysis in the Oregon-ADHD-1000 data set, 75% of participants were randomly selected for training the classifiers (and to estimate performance using 5-fold cross validation), and the remaining 25% were used to validate classifier performance in a hold-out sample. Table 1 reveals no statistically significant differences between the training and test data sets in terms of demographic or clinical features. Differences between the Oregon and Michigan cohorts are notable in regard to generalizability and are discussed later.

**Table 1.** Overview of samples used in the training and testing of the classifiers. ^‡^The percentage of subjects with any current diagnosis of anxiety, mood, or disruptive (ODD/CD) disorder. *P-values <0.05 for statistical tests comparing the Michigan-1000 cohort to the entire Oregon-1000 cohort. P-ADHDRS=parent ADHD Rating Scale, T-ADHDRS=teacher ADHD Rating Scale.

|  | Oregon-1000 |  | Michigan-1000 |
| --- | --- | --- | --- |
|  | Training Set | Test Set |  |
| Total N | 1067 | 356 | 1057 |
| ADHD cases | 543 (50.9%) | 181 (50.8%) | 458 (43.3%)* |
| Controls | 318 (29.8%) | 104 (29.2%) | 472 (44.7%)* |
| Other | 206 (19.3%) | 71 (19.9%) | 127 (12.0%)* |
| % Males | 60.1 | 62.4 | 56.5* |
| Mean Age (SD) | 9.4 (1.5) | 9.4 (1.7) | 12.5 (3.1)* |
| % White / Caucasian | 79.9% | 77.0% | 72.5%* |
| P-ADHDRS Int | 62.3 (16.5) | 61.3 (16) | 55.0 (14.7)* |
| P-ADHDRS Hyp | 59.2 (15.9) | 58.6 (15.9) | 51.6 (12.9)* |
| P-Conners Int | 64.5 (16.4) | 64.3 (15.8) | 60.5 (14.8)* |
| P-Conners Hyp | 63.0 (17.3) | 62.2 (16.9) | 59.0 (15.4)* |
| T-ADHDRS Int | 51.8 (10.1) | 52.1 (10.1) | 48.8 (9.8)* |
| T-ADHDRS Hyp | 51.0 (9.3) | 51.3 (9.8) | 48.3 (8.2)* |

As an initial, qualitative examination of how well the predictive features were able to separate diagnostic classes, we visualized the Oregon training set by performing t-SNE (van der Maaten and Hinton 2008), a non-linear dimensionality reduction technique, using the same subsets of predictive features as will be used in our multi-stage classifier (parent-reported symptom, teacher-reported symptoms, and cognitive measures). Since t-SNE maps the distribution of similarities between pairs of high-dimensional entities to a distribution in a low-dimensional space (here two dimensions), it can be useful for assessing the structure (e.g., presence of clusters) in high-dimensional data. Supplemental Figure S1 shows that parent and teacher ratings alone provided initial meaningful separation between ADHD and non-ADHD groups or clusters. As might be expected, those participants who the best-estimate procedure considered as either subthreshold or other appeared more difficult to classify.

### Comparison of Classifier Performances on Predicting Best-estimate Diagnosis

The 5-fold cross-validation and test-set accuracies for all classifiers evaluated on the Oregon cohort are reported in **Table 2**. The table also includes accuracies on the subset of participants given a high-confidence prediction (class probability >0.9). Additional performance measures for all classifiers are reported in Supplemental Tables S4 and S5. All classifiers were trained, using all discretized predictive features, to distinguish ADHD cases from controls (including *subthreshold* and *other* to partially mimic the difficulty in clinical diagnosis). Receiver operating characteristic (ROC) curves for all classifiers are shown in Supplemental Figure S2.

**Table 2.** Mean 5-fold cross-validation (CV) and test-set accuracies for the classifiers predicting best-estimate team diagnoses in the Oregon cohort. The classifiers included all discretized parent, teacher, and cognitive predictive features. The optimal hyperparameters used for each classifier are given in Supplemental Table S3. The test set is the Oregon 25% hold out sample (N=356). *Accuracies on the subset of the test set with high-confidence predictions (predicted class probability >0.9); the number of high-confidence predictions and the corresponding percentage of the test set are given in parentheses. SD=standard deviation, LR=logistic regression, DT=decision tree, TAN=tree-augmented naïve Bayes, RF=random forest, SVM=support vector machine, GBDT=gradient-boosted decision trees.

| Classifier | 5-fold CV (SD) | Test Set | Test Set, High-confidence* |
| --- | --- | --- | --- |
| LR-unregularized | 0.851 (0.011) | 0.882 | 0.964 (N=223; 63%) |
| DT-simple | 0.840 (0.011) | 0.860 | 0.936 (N=251; 71%) |
| TAN-stage4 | 0.868 (0.014) | 0.874 | 0.908 (N=326; 92%) |
| TAN-earliest | 0.869 (0.015) | 0.860 | 0.870 (N=339; 95%) |
| LR | 0.859 (0.012) | 0.876 | 0.971 (N=208; 58%) |
| DT | 0.870 (0.015) | 0.882 | 0.953 (N=191; 54%) |
| RF | 0.884 (0.005) | 0.910 | 0.969 (N=191; 54%) |
| SVM | 0.877 (0.009) | 0.924 | 0.975 (N=237; 67%) |
| GBDT | 0.890 (0.004) | 0.930 | 0.974 (N=229; 64%) |
| Ensemble | 0.875 (0.013) | 0.899 | 0.968 (N=219; 62%) |

All classifiers were able to predict the best-estimate team diagnosis with 5-fold cross-validation accuracy >85% (base rate ∼51%), except for the simple 2-level decision tree. Gradient boosted decision trees performed significantly better (89% mean 5-fold cross-validation accuracy) than both “simpler” methods, unregularized logistic regression (85%) and the 2-level decision tree (84%). However, there were no significant differences among the more advanced classifiers (Supplemental Table S6). Cross-validation performance was representative of performance on the held-out Oregon test set, with test set accuracies ranging from 87.4% for TAN to 93% for gradient boosted decision trees.

For the TAN classifier, using the earliest high-confidence prediction (“TAN-earliest” in Table 2; Figure 1B) resulted in very similar overall performance compared to using the prediction after all 4 stages. However, the accuracy of high-confidence predictions in the test set was slightly lower for TAN-earliest.

#### Sensitivity analysis

The performance of all classifiers, except for TAN, when using the continuous predictive measures, as well as the performance for different discretization methods, is reported in the Supplemental Materials (Supplemental Tables S7 and S8). We saw no significant differences in cross-validation accuracy when using discretized predictors vs. continuous predictors for any of the classification methods.

### Classifier Performance Predicting KSADS Diagnosis

Because the best-estimate team used some of the same predictive features as used in the classifiers, we also evaluated the ability of the classifiers to predict ADHD status defined using KSADS measures (i.e., the labels were determined independently from all predictive features used in the classifier). Agreement between best-estimate team and KSADS ADHD versus non-ADHD status was 85% (see Supplemental Table S9).

The performance of classifiers trained to predict KSADS labels in the Oregon cohort is reported in Supplemental Tables S10, S11, and S12. Overall, classification accuracy of KSADS diagnoses was slightly lower than seen for the best-estimate team diagnoses. All classifiers achieved mean 5-fold cross-validation accuracy >82%, with the TAN classifier showing the highest cross-validation accuracy (84.8%) and regularized logistic regression the lowest (82.8%). There were no significant differences in mean 5-fold cross-validation accuracies across the classification methods (Supplemental Table S13). And again, the 5-fold cross-validation accuracies were fairly representative of performance on the test set, which ranged from 81.5% accuracy for the decision tree to 84.8% accuracy for gradient-boosted decision trees.

### The Value of Confidence Thresholds to Determine Classifications

The multi-stage TAN approach allows for classifications to be assigned based on only a subset of the data, if that data produces a high-confidence classification (i.e., a class probability >0.9 in this case). Given the cost of gathering diagnostic data, particularly from laboratory tests, this approach could potentially save significant resources without sacrificing classification performance. Resources could be saved for difficult-to-classify cases.

We examined the proportion of subjects who could be classified with high confidence at each stage of the TAN classifier, as well as the accuracy of these high-confidence classifications. **Figure 2** shows that a significant portion of the test-set subjects (312/356, 87.6%) are classified with high-confidence (class probability >0.9) using only parent and teacher reported symptom scales (i.e., stages 1-3). The accuracy of these high-confidence classifications at stage 3 was 88.8% in the Oregon hold-out test set.

**Figure 2.**
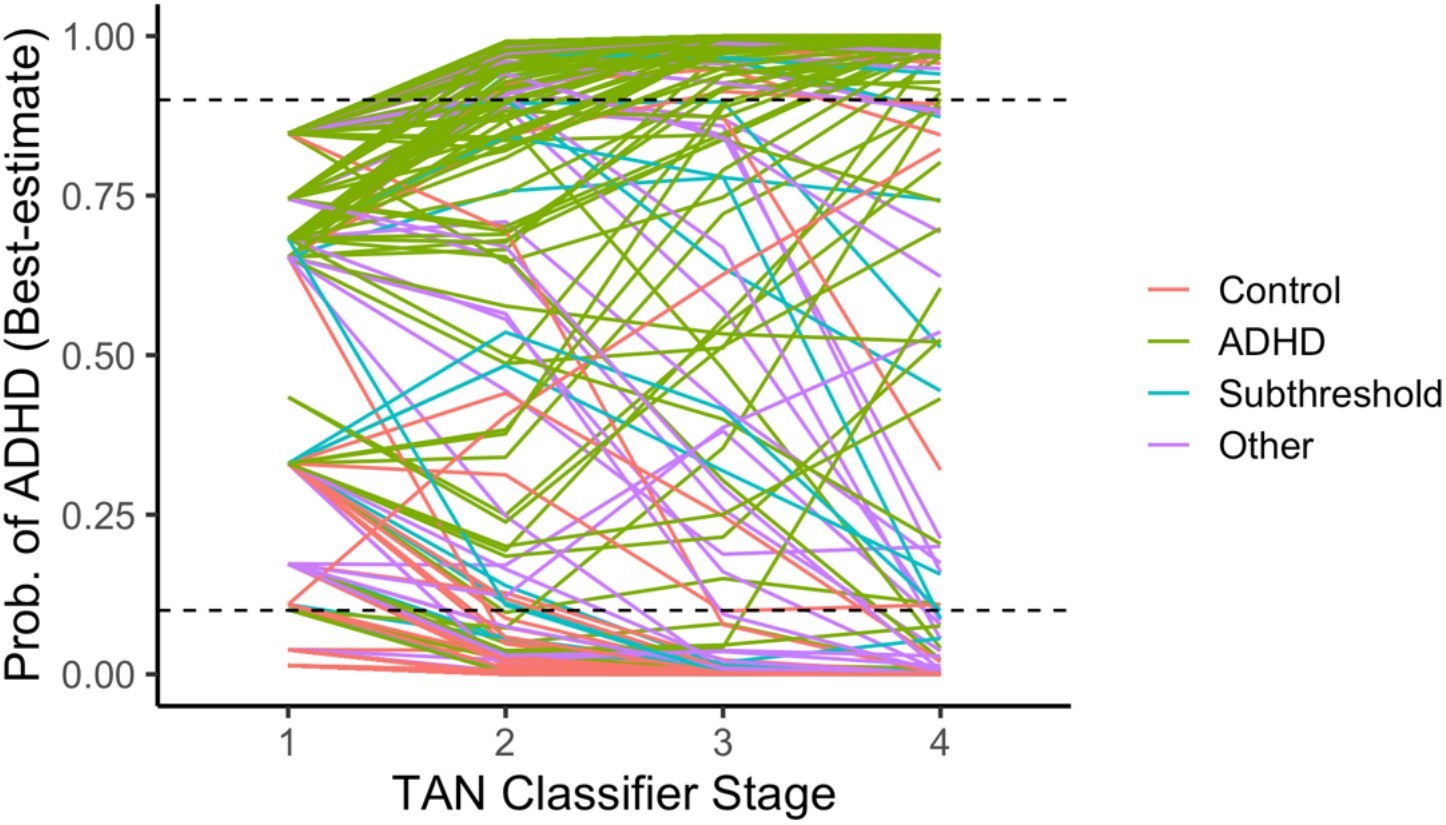
Predicted probability of an ADHD diagnosis (best-estimate team), at each stage of the TAN classifier for all participants in the Oregon test set. Most ADHD cases converge correctly towards a probability of ADHD of 1.0 across the 4 stages. Likewise, most controls converge correctly towards a probability of ADHD of 0. The majority of participants are predicted with high confidence (i.e., a predicted probability >0.9 or <0.1; dashed horizontal lines), by stage 3. Yet a small subset of participants had unstable predictions (large changes in predicted probability in subsequent stages)—these are significantly more likely to be “other” or subthreshold individuals (see Supplemental Materials). Stage 1 = parent ADHD Rating Scale, Stage 2 = parent Conners, Stage 3 = teacher ADHD Rating Scale, and Stage 4 = cognitive measures.

For the remaining 44 test-set subjects with low-confidence classifications (class probability <0.9) after stage 3 of the TAN classifier, the addition of data from cognitive tests resulted in: (1) a class probability increase for N=26 (22 of which became high-confidence; 20 classified correctly), (2) a class probability decrease for N=18, and (3) a change in predicted class for N=10 (including 9 correctly classified; 4 with high confidence). Overall, when cognitive measures were included in the classifier (stage 4), the TAN classifier predicted 92% (326/356) of the Oregon test set with high confidence. The accuracy of these high-confidence predictions was 90.8%. The misclassified cases are discussed further below.

As expected, the accuracy of high-confidence predictions is significantly better than lower-confidence predictions. Therefore, we examined how often each classifier provided a high-confidence prediction (predicted class probability >0.9). As shown in the parenthetical numbers in the right hand column of **Table 2**, the TAN-stage4 classifier strongly outperformed other methods in providing the most high-confidence predictions (92% of the test set; accuracy of 91% on this subset of high-confidence, test-set predictions), likely due to its ability to incorporate prior probabilities in subsequent stages of the classifier. The proportion of high-confidence classifications for other methods was far lower (Table 2).

In other words, the TAN classifier was able to provide high-confidence predictions for a higher percentage of the sample than the other methods. These results translate into the TAN classifier correctly classifying an additional 61 participants (17.1% of Oregon test set; 21 true positive ADHD cases) with high confidence, and incorrectly classifying an additional 14 participants (3.9% of Oregon test set; 7 false positives) with high-confidence, compared to the classifier with the next highest number of high-confidence classifications (DT-simple).

At the same time, small number of participants in the Oregon cohort test set were misclassified with high confidence by the TAN classified (N=30; 8.4% of the test set). Of the 18 high-confidence false positives, 16 were either subthreshold (N=5) or other (N=11), highlighting the diagnostic difficulty of those cases. Of the 12 high-confidence false negatives, 10 had scores of 1=“minimal” on the KSADS for ADHD current impairment (scores range from 0=“none” to 3=“severe”; the other two had scores of 2=“moderate”), and all had a parent-reported SDQ impact score of 0 (0=“normal”, 1=“borderline”, 2=“abnormal”). Thus, addition of a separate impairment rating in the algorithm would improve accuracy.

Class probabilities, at all stages of the TAN classifier, for all subjects in the Oregon test set are shown in **Figure 2**. It shows that (1) most participants were classified with high confidence (probability of ADHD ≥0.9) at Stage 3, prior to incorporating the cognitive measures; (2) a small subset were never classified with high confidence, even when all predictors were in the classifier; and (3) some participants have conflicting data at different stages (their class probabilities are highly variable, or “unstable”, across the stages).

The online supplement provides a sensitivity analysis regarding the classification difficulty of the sub-threshold and “other” cases.

### Sensitivity analysis: Impact of “other” or borderline classified participants

Given the diagnostic uncertainty implied by the subthreshold and other categories, we investigated the effect of re-labeling or excluding these subjects from the data used to train the classifiers. The performance of classifiers trained on the full training data, with subthreshold and other cases all labeled as non-ADHD, is referred to as Training Scheme A. It was compared to the performance of two other training schemes. The results are displayed in Table 3. In Training Scheme B, the full training data set was used, but subthreshold cases were selected and re-labeled as ADHD cases. In Training Scheme C, all subthreshold and other cases were excluded from the training set. Scheme B led to reduced accuracy (Table 3). Scheme C resulted, not surprisingly, in slightly increased sensitivity (test set sensitivity 0.939 vs. 0.917), but reduced specificity (test set specificity 0.760 vs. 0.829).

**Table 3.** Comparison of TAN classifier performance with three different training schemes. Training scheme A includes all training set subjects, with subthreshold and not-clean controls labeled as controls; Training scheme B re-labels subthreshold subjects as ADHD cases; and Training scheme C excludes subthreshold and not-clean control subjects from the training set (but they are retained in the test set). *CV performance measures are the average of measures across all 5-folds of the training set (average N=213). The full independent test set consists of N=356 subjects. Performance is also reported for only those subjects given a high-confidence (P 2 0.9) prediction (N=326, 323, and 336 for the three training schemes, respectively). **Bolded** values are the highest among the three training schemes for the test set, while *italicized* values are the highest among the three training schemes for CV. PPV_X_=positive predictive value assuming X% prevalence of ADHD.

|  | Scheme A |  | Scheme B |  | Scheme C |  |
| --- | --- | --- | --- | --- | --- | --- |
| Full CV/Test Set* | 5-fold CV | Test | 5-fold CV | Test | 5-fold CV | Test |
| Accuracy | *0.868* | **0.874** | 0.866 | 0.854 | 0.857 | 0.851 |
| Sensitivity | 0.912 | 0.917 | 0.900 | 0.900 | *0.959* | **0.939** |
| Specificity | *0.823* | **0.829** | 0.818 | 0.805 | 0.750 | 0.760 |
| PPV<sub>5</sub> | *0.213* | **0.220** | 0.206 | 0.194 | 0.168 | 0.171 |
| PPV<sub>50</sub> | *0.837* | **0.843** | 0.832 | 0.820 | 0.793 | 0.796 |
| AUC-ROC | *0.947* | **0.942** | 0.937 | 0.921 | 0.910 | 0.915 |
| High-confidence Predictions Only | N=972 | N=326 | N=986 | N=323 | N=1017 | N=336 |
| Accuracy | *0.903* | **0.908** | 0.886 | 0.885 | 0.870 | 0.872 |
| Sensitivity | 0.943 | 0.930 | 0.923 | 0.910 | *0.973* | **0.960** |
| Specificity | *0.862* | **0.884** | 0.835 | 0.851 | 0.761 | 0.778 |
| PPV<sub>5</sub> | *0.264* | **0.296** | 0.227 | 0.243 | 0.176 | 0.185 |
| PPV<sub>50</sub> | *0.872* | **0.889** | 0.848 | 0.859 | 0.803 | 0.812 |
| AUC-ROC | *0.959* | **0.950** | 0.949 | 0.934 | 0.913 | 0.918 |

### Generalization to the Michigan-ADHD-1000 Cohort

Finally, we evaluated whether a classifier trained on the entire Oregon data set would perform well on the independent Michigan cohort. As in the primary analyses done in the Oregon cohort, subjects categorized as subthreshold or other were labeled as controls for the purposes of training the classifiers in the full Oregon cohort. Because a different version of the Conners rating scale (with fewer subscales) was used in the Michigan cohort (due to its earlier period of collection), new classifiers were trained on the Oregon data excluding the subscales that were not available in the Michigan data.

Several differences between the two cohorts were expected to challenge the generalizability of the model but also provided an important opportunity to examine real-world generalizability/reproducibility questions (Table 1). Notably, the Michigan cohort included a significantly wider age range, and higher average age (12.5 vs. 9.4 years), a lower proportion of ADHD cases, a lower proportion of males, and higher proportion of non-white individuals.

Finally, the overall distribution of symptom measures is shifted further into the clinical range in the Michigan cohort compared to the Oregon cohort—this difference remains when examining ADHD cases only (data not shown), suggesting the Michigan cohort is a somewhat more clinically severe ADHD sample.

Thus, it was encouraging that despite these differences, most classifiers showed impressive generalizability to the Michigan cohort, for classifying best-estimate team diagnosis. **Table 4** shows these results. The TAN, RF, SVM, and GBDT methods were all able to classify ADHD and non-ADHD (including “other” and subthreshold) with >79% accuracy. Again, there appears to be a slight advantage to the more advanced classifiers, with GBDT performing best (82% accuracy) and the 2-level decision tree worst (76% accuracy) in the Michigan cohort. As seen in the Oregon cohort, the TAN provided considerably more high-confidence predictions (91%) than other methods (41%-69%).

**Table 4.** Accuracy of the classifiers predicting best-estimate team diagnoses in the Michigan cohort. Also show (last column) are the accuracies for the subset of Michigan cohort subjects given high-confidence predictions (predicted class probability >0.9); the number of high-confidence predictions and the corresponding percentage of the Michigan cohort are given in parentheses. The classifiers were trained on data from the entire Oregon cohort, using all available (discretized) parent, teacher, and cognitive predictive features.

| Classifier | Training Set (Oregon; N=1423) | Test Set (Michigan; N=1057) | Test Set (Michigan; High-confidence) |
| --- | --- | --- | --- |
| LR-unregularized | 0.878 | 0.779 | 0.895 (N=636; 60%) |
| DT-simple | 0.850 | 0.761 | 0.879 (N=725; 69%) |
| TAN-stage4 | 0.877 | 0.792 | 0.820 (N=957; 91%) |
| LR | 0.874 | 0.785 | 0.906 (N=585; 55%) |
| DT | 0.878 | 0.770 | 0.917 (N=435; 41%) |
| RF | 0.940 | 0.807 | 0.932 (N=474; 45%) |
| SVM | 0.916 | 0.799 | 0.902 (N=673; 64%) |
| GBDT | 0.909 | 0.822 | 0.909 (N=635; 60%) |
| Ensemble | 0.919 | 0.807 | 0.916 (N=585; 55%) |

Again, we observed slightly poorer performance across all methods when classifying KSADS diagnoses. The TAN performed best in that case, with an accuracy of 78.9% (Supplemental Tables S15 – S18). Overall, generalizability to a new cohort was supportive of the potential for these algorithms to achieve useful accuracy.

## DISCUSSION

Classification of gold-standard research diagnosis of ADHD was achieved with fairly high accuracy overall (>85%), including generally with only parent and teacher total rating scale scores. In terms of classification accuracy, advanced methods appear to provided only incremental improvement over unregularized logistic regression and a simple (2-level) decision tree. However, this slight benefit diminished when predicting KSADS diagnosis, and when examining generalizability to a completely independent cohort. Likewise, overall performance was statistically similar across the more advanced classifiers.

At same time, however, because the TAN classifier was readily able to incorporate prior probabilities from early-stage predictions into the later-stages, it provided high-confidence predictions for many more subjects than other classifiers. This may be a particularly advantageous feature of the TAN, given that prediction confidence will be an important factor for determining clinical utility. In the Oregon held-out test set (N=356), the high-confidence predictions from the TAN classifier resulted in an additional 40 high-confidence true negatives, 21 high-confidence true positives, 7 high-confidence false positives, and 7 high-confidence false negatives, compared to other methods. In other words, an additional 17% of the sample was correctly classified with high confidence, with only a 4% increase in high-confidence misclassifications.

Although overall accuracies were comparable across the more advanced classification methods, the Bayesian nature of the TAN classifier may align better with clinical decision-making processes, increasing interpretability and potential clinical utility. The multi-stage TAN classifier demonstrated performance comparable to other advanced methods, generalized well, and provided evidence that a large majority of subjects can be classified accurately with only low-cost parent and teacher symptom reports, even with very difficult borderline cases in the sample. The addition of data from laboratory cognitive tests provided a benefit, in terms of prediction confidence, for only a small subset of cases. For instance, of the participants with low-confidence classifications based on parent and teacher measures alone, half (or 6% of the total test sample) benefited from further data from cognitive tests. This was largely consistent with results from a smaller study of younger children (Öztekin et al. 2021). Identifying those subjects who will benefit from further assessment (of all kinds) should be a priority for future work.

The posterior class probability is an intuitive way to judge the confidence of a classification. Here we used a class probability cutoff of 0.9 to define a “high-confidence” prediction, but there is no firm guidance and this criterion was ultimately arbitrary. The optimal threshold to use for a particular classifier and outcome is still an open question, and should be studied further with the potential costs of misclassification in mind.

Another conclusion suggested by our findings is that a training data set representing the full spectrum of ADHD presentations, including subthreshold cases and non-ADHD subjects with other psychiatric conditions, may be important for reducing false-positive classifications. A full spectrum provides better granularity which might help better defined decision boundaries for the ADHD class. This finding should be confirmed with additional data, given that the impact of uncertain diagnoses may change as the size of training sets increase.

The current study has a number of strengths, including: a larger sample size than most prior attempts at single case prediction in ADHD using machine learning, the inclusion of a completely independent validation cohort for independent generalizability (rare in this literature), the inclusion of difficult borderline diagnostic cases, a competitive modeling approach, and the examination of the relative importance of predictive measures in a multi-stage classification approach. Each of these provides a valuable contribution to the literature and in general, results are promising.

Certain limitations do bear mention. First, despite the large samples used for training and evaluating the classifiers, and the availability of a generalizability cohort, substantial further work would be needed to evaluate generalizability and site-specific algorithms for clinical use. Second, the large amount of missing data among the cognitive measures, and the confound between diagnostic category and availability of these data, limited our ability to make strong conclusions about the utility of those measures. Finally, our reliance on a method (TAN) that required discretized measures could have resulted in loss of information, and therefore could have hurt performance—although our sensitivity analysis did not suggest this was the case.

That symptom thresholds are typically used for diagnosis suggests that discretization of measures is appropriate (and possibly beneficial, due to removing redundant information) for the current application. The sensitivity analyses conducted, regarding missing data and discretization, lend confidence to our conclusions.

In conclusion, the findings reported here point to the potential utility of machine learning to aid in fast, low-cost diagnosis of ADHD in children. Future research should focus on (1) further validation of classifiers in larger and more diverse samples, including investigating predictive ability across a wider age range, (2) identification of difficult-to-classify subjects and those who will benefit from additional, more costly assessment (including, e.g., genetic risk scores), and (3) evaluation of the cost/benefit of prediction confidence thresholds that could guide clinical deployment of advanced classification algorithms.

## Data Availability

Data from both the Oregon-ADHD-1000 (collection 2857) and Michigan-ADHD-1000 (collection 3753; in progress) are being made available on the NIMH Data Archive.

https://nda.nih.gov/edit_collection.html?id=2857

https://nda.nih.gov/edit_collection.html?id=3753

## ACKNOWLEDGEMENTS

This work was supported by funding from the National Institute of Mental Health (R37MH059105).

## REFERENCES

1 Abu-Mostafa, Yaser S., Malik Magdon-Ismail, and Hsuan-Tien Lin. 2012. Learning From Data. AMLBook.

2 Adler, Lenard, David Shaw, David Sitt, Erica Maya, and Melinda Ippolito Morrill. 2009. “Issues in the Diagnosis and Treatment of Adults ADHD by Primary Care Physicians.” Primary Psychiatry 16 (5): 57–63.

3 APA. 2013. Diagnostic and Statistical Manual of Mental Disorders: DSM-5™, 5th Ed. Diagnostic and Statistical Manual of Mental Disorders: DSM-5™, 5th Ed. Arlington, VA, US: American Psychiatric Publishing, Inc. https://doi.org/10.1176/appi.books.9780890425596.

4 Bishop, Christopher M. 2006. Pattern Recognition and Machine Learning. Information Science and Statistics. New York: Springer-Verlag. https://www.springer.com/gp/book/9780387310732.

5 Bledsoe, Jesse C., Danica Xiao, Art Chaovalitwongse, Sonya Mehta, Thomas J. Grabowski, Margaret Semrud-Clikeman, Steven Pliszka, and David Breiger. 2016. “Diagnostic Classification of ADHD Versus Control: Support Vector Machine Classification Using Brief Neuropsychological Assessment.” Journal of Attention Disorders, May, 1087054716649666. https://doi.org/10.1177/1087054716649666.

6 Cella, David, Susan Yount, Nan Rothrock, Richard Gershon, Karon Cook, Bryce Reeve, Deborah Ader, et al. 2007. “The Patient-Reported Outcomes Measurement Information System (PROMIS): Progress of an NIH Roadmap Cooperative Group during Its First Two Years.” Medical Care 45 (5 Suppl 1): S3–11. https://doi.org/10.1097/01.mlr.0000258615.42478.55.

7 Christiansen, Hanna, Mira-Lynn Chavanon, Oliver Hirsch, Martin H. Schmidt, Christian Meyer, Astrid Müller, Hans-Juergen Rumpf, Ilya Grigorev, and Alexander Hoffmann. 2020. “Use of Machine Learning to Classify Adult ADHD and Other Conditions Based on the Conners’ Adult ADHD Rating Scales.” Scientific Reports 10 (1): 18871. https://doi.org/10.1038/s41598-020-75868-y.

8 Committee on Quality Improvement, Subcommittee on Attention-Deficit/Hyperactivity Disorder. 2000. “Clinical Practice Guideline: Diagnosis and Evaluation of the Child with Attention-Deficit/Hyperactivity Disorder. American Academy of Pediatrics.” Pediatrics 105 (5): 1158–70. https://doi.org/10.1542/peds.105.5.1158.

9 Conners, C. Keith. 2003. Connors’ Rating Scales: Revised Technical Manual. New York, NY: Multi-Health Systems.

10 Conners, C. Keith. 2008. Conners 3rd Edition Manual. Toronto, Ontario, Canada: Multi-Health Systems.

11 Conners, C. Keith, Gill Sitarenios, James D. A. Parker, and Jeffery N. Epstein. 1998. “The Revised Conners’ Parent Rating Scale (CPRS-R): Factor Structure, Reliability, and Criterion Validity.” Journal of Abnormal Child Psychology 26 (4): 257–68. https://doi.org/10.1023/A:1022602400621.

12 Costello, E. Jane, Jian-ping He, Nancy A. Sampson, Ronald C. Kessler, and Kathleen Ries Merikangas. 2014. “Services for Adolescents with Psychiatric Disorders: 12-Month Data from the National Comorbidity Survey-Adolescent.” Psychiatric Services (Washington, D.C.) 65 (3): 359–66. https://doi.org/10.1176/appi.ps.201100518.

13 De Luca, Cinzia R., Stephen J. Wood, Vicki Anderson, Jo-Anne Buchanan, Tina M. Proffitt, Kate Mahony, and Christos Pantelis. 2003. “Normative Data From the Cantab. I: Development of Executive Function Over the Lifespan.” Journal of Clinical and Experimental Neuropsychology 25 (2): 242–54. https://doi.org/10.1076/jcen.25.2.242.13639.

14 Delis, Dean C., Edith Kaplan, and Joel H. Kramer. 2001. Delis-Kaplan Executive Function System. Psychological Corporation.

15 Dietterich, Thomas G. 1998. “Approximate Statistical Tests for Comparing Supervised Classification Learning Algorithms.” Neural Computation 10 (7): 1895–1923. https://doi.org/10.1162/089976698300017197.

16 Duda, M., R. Ma, N. Haber, and D. P. Wall. 2016. “Use of Machine Learning for Behavioral Distinction of Autism and ADHD.” Translational Psychiatry 6 (2): e732–e732. https://doi.org/10.1038/tp.2015.221.

17 DuPaul, G, T Power, A Anastopoulos, and R Reid. 1998. ADHD Rating Scale—IV: Checklists, Norms, and Clinical Interpretation. NY, NY: Guilford Press.

18 Dvorsky, Melissa R., Joshua M. Langberg, Stephen J. Molitor, and Elizaveta Bourchtein. 2016. “Clinical Utility and Predictive Validity of Parent and College Student Symptom Ratings in Predicting an ADHD Diagnosis.” Journal of Clinical Psychology 72 (4): 401–18. https://doi.org/10.1002/jclp.22268.

19 Faraone, Stephen V., Thomas J. Spencer, C. Brendan Montano, and Joseph Biederman. 2004. “Attention-Deficit/Hyperactivity Disorder in Adults: A Survey of Current Practice in Psychiatry and Primary Care.” Archives of Internal Medicine 164 (11): 1221–26. https://doi.org/10.1001/archinte.164.11.1221.

20 Gibbons, Robert D., David J. Kupfer, Ellen Frank, Benjamin B. Lahey, Brandie A. George-Milford, Candice L. Biernesser, Giovanna Porta, Tara L. Moore, Jong Bae Kim, and David A. Brent. 2020. “Computerized Adaptive Tests for Rapid and Accurate Assessment of Psychopathology Dimensions in Youth.” Journal of the American Academy of Child and Adolescent Psychiatry 59 (11): 1264–73. https://doi.org/10.1016/j.jaac.2019.08.009.

21 Gibbons, Robert D., David J. Weiss, Ellen Frank, and David Kupfer. 2016. “Computerized Adaptive Diagnosis and Testing of Mental Health Disorders.” Annual Review of Clinical Psychology 12: 83–104. https://doi.org/10.1146/annurev-clinpsy-021815-093634.

22 Gill, Christopher J, Lora Sabin, and Christopher H Schmid. 2005. “Why Clinicians Are Natural Bayesians.” BMJ : British Medical Journal 330 (7499): 1080–83.

23 Goodfellow, Ian, Yoshua Bengio, and Aaron Courville. 2016. Deep Learning. MIT Press.

24 Hall, Charlotte L., Althea Z. Valentine, Madeleine J. Groom, Gemma M. Walker, Kapil Sayal, David Daley, and Chris Hollis. 2016. “The Clinical Utility of the Continuous Performance Test and Objective Measures of Activity for Diagnosing and Monitoring ADHD in Children: A Systematic Review.” European Child & Adolescent Psychiatry 25 (7): 677–99. https://doi.org/10.1007/s00787-015-0798-x.

25 Irwin, Debra E., Brian Stucky, Michelle M. Langer, David Thissen, Esi Morgan Dewitt, Jin-Shei Lai, James W. Varni, Karin Yeatts, and Darren A. DeWalt. 2010. “An Item Response Analysis of the Pediatric PROMIS Anxiety and Depressive Symptoms Scales.” Quality of Life Research: An International Journal of Quality of Life Aspects of Treatment, Care and Rehabilitation 19 (4): 595–607. https://doi.org/10.1007/s11136-010-9619-3.

26 Jerez José M., Ignacio Molina, Pedro J. García-Laencina, Emilio Alba, Nuria Ribelles, Miguel Martín, and Leonardo Franco. 2010. “Missing Data Imputation Using Statistical and Machine Learning Methods in a Real Breast Cancer Problem.” Artificial Intelligence in Medicine 50 (2): 105–15. https://doi.org/10.1016/j.artmed.2010.05.002.

27 Karalunas, Sarah L., Hanna C. Gustafsson, Nathan F. Dieckmann, Jessica Tipsord, Suzanne H. Mitchell, and Joel T. Nigg. 2017. “Heterogeneity in Development of Aspects of Working Memory Predicts Longitudinal Attention Deficit Hyperactivity Disorder Symptom Change.” Journal of Abnormal Psychology 126 (6): 774–92. https://doi.org/10.1037/abn0000292.

28 Loskutova, Natalia Y., Cory B. Lutgen, Elisabeth F. Callen, Melissa K. Filippi, and Elise A. Robertson. 2021. “Evaluating a Web-Based Adult ADHD Toolkit for Primary Care Clinicians.” Journal of the American Board of Family Medicine: JABFM 34 (4): 741–52. https://doi.org/10.3122/jabfm.2021.04.200606.

29 Maaten, L. J. P. van der, and G. E. Hinton. 2008. “Visualizing High-Dimensional Data Using t-SNE.” Journal of Machine Learning Research 9 (nov): 2579–2605.

30 Marek, Scott, Brenden Tervo-Clemmens, Finnegan J. Calabro, David F. Montez, Benjamin P. Kay, Alexander S. Hatoum, Meghan Rose Donohue, et al. 2020. “Towards Reproducible Brain-Wide Association Studies.” BioRxiv, August, 2020.08.21.257758. https://doi.org/10.1101/2020.08.21.257758.

31 Martin, Alicia R., Mark J. Daly, Elise B. Robinson, Steven E. Hyman, and Benjamin M. Neale. 2019. “Predicting Polygenic Risk of Psychiatric Disorders.” Biological Psychiatry 86 (2): 97–109. https://doi.org/10.1016/j.biopsych.2018.12.015.

32 Martin, Alicia R., Masahiro Kanai, Yoichiro Kamatani, Yukinori Okada, Benjamin M. Neale, and Mark J. Daly. 2019. “Current Clinical Use of Polygenic Scores Will Risk Exacerbating Health Disparities.” Nature Genetics 51 (4): 584–91. https://doi.org/10.1038/s41588-019-0379-x.

33 Massuti, Rafael, Carlos Renato Moreira-Maia, Fausto Campani, Márcio Sônego, Julia Amaro, Gláucia Chiyoko Akutagava-Martins, Luca Tessari, Guilherme V. Polanczyk, Samuele Cortese, and Luis Augusto Rohde. 2021. “Assessing Undertreatment and Overtreatment/Misuse of ADHD Medications in Children and Adolescents across Continents: A Systematic Review and Meta-Analysis.” Neuroscience and Biobehavioral Reviews 128 (September): 64–73. https://doi.org/10.1016/j.neubiorev.2021.06.001.

34 Mihaljević, Bojan, Concha Bielza, and Pedro Larrañaga. 2018. “Bnclassify: Learning Bayesian Network Classifiers.” The R Journal 10 (2): 455–68.

35 Mooney, Michael A., Priya Bhatt, Robert J. M. Hermosillo, Peter Ryabinin, Molly Nikolas, Stephen V. Faraone, Damien A. Fair, Beth Wilmot, and Joel T. Nigg. 2021. “Smaller Total Brain Volume but Not Subcortical Structure Volume Related to Common Genetic Risk for ADHD.” Psychological Medicine 51 (8): 1279–88. https://doi.org/10.1017/S0033291719004148.

36 Mooney, Michael A., Peter Ryabinin, Beth Wilmot, Priya Bhatt, Jonathan Mill, and Joel T. Nigg. 2020. “Large Epigenome-Wide Association Study of Childhood ADHD Identifies Peripheral DNA Methylation Associated with Disease and Polygenic Risk Burden.” Translational Psychiatry 10 (1): 1–12. https://doi.org/10.1038/s41398-020-0710-4.

37 Mueller, Andreas, Gian Candrian, Juri D. Kropotov, Valery A. Ponomarev, and Gian-Marco Baschera. 2010. “Classification of ADHD Patients on the Basis of Independent ERP Components Using a Machine Learning System.” Nonlinear Biomedical Physics 4 (1): S1. https://doi.org/10.1186/1753-4631-4-S1-S1.

38 National Guideline Centre (UK). 2018. Attention Deficit Hyperactivity Disorder: Diagnosis and Management. National Institute for Health and Care Excellence: Clinical Guidelines. London: National Institute for Health and Care Excellence (UK). http://www.ncbi.nlm.nih.gov/books/NBK493361/.

39 Nigg, Joel T. 1999. “The ADHD Response-Inhibition Deficit as Measured by the Stop Task: Replication with DSM-IV Combined Type, Extension, and Qualification.” Journal of Abnormal Child Psychology 27 (5): 393–402. https://doi.org/10.1023/a:1021980002473.

40 Nigg, Joel T., Hanna C. Gustafsson, Sarah L. Karalunas, Peter Ryabinin, Shannon K. McWeeney, Stephen V. Faraone, Michael A. Mooney, Damien A. Fair, and Beth Wilmot. 2018. “Working Memory and Vigilance as Multivariate Endophenotypes Related to Common Genetic Risk for Attention-Deficit/Hyperactivity Disorder.” Journal of the American Academy of Child and Adolescent Psychiatry 57 (3): 175–82. https://doi.org/10.1016/j.jaac.2017.12.013.

41 Nigg, Joel T., Sarah L. Karalunas, Hanna C. Gustafsson, Priya Bhatt, Peter Ryabinin, Michael A. Mooney, Stephen V. Faraone, Damien A. Fair, and Beth Wilmot. 2020. “Evaluating Chronic Emotional Dysregulation and Irritability in Relation to ADHD and Depression Genetic Risk in Children with ADHD.” Journal of Child Psychology and Psychiatry 61 (2): 205–14. https://doi.org/10.1111/jcpp.13132.

42 Nikolas, Molly A., Paul Marshall, and James B. Hoelzle. 2019. “The Role of Neurocognitive Tests in the Assessment of Adult Attention-Deficit/Hyperactivity Disorder.” Psychological Assessment 31 (5): 685–98. https://doi.org/10.1037/pas0000688.

43 Nikolas, Molly A., and Joel T. Nigg. 2013. “Neuropsychological Performance and Attention-Deficit Hyperactivity Disorder Subtypes and Symptom Dimensions.” Neuropsychology 27 (1): 107–20. https://doi.org/10.1037/a0030685.

44 Nikolas, Molly A., and Joel T. Nigg. 2015. “Moderators of Neuropsychological Mechanism in Attention-Deficit Hyperactivity Disorder.” Journal of Abnormal Child Psychology 43 (2): 271–81. https://doi.org/10.1007/s10802-014-9904-7.

45 Olson, Randal S., William La Cava, Zairah Mustahsan, Akshay Varik, and Jason H. Moore. 2018. “Data-Driven Advice for Applying Machine Learning to Bioinformatics Problems.” Pacific Symposium on Biocomputing. Pacific Symposium on Biocomputing 23: 192–203.

46 Öztekin, Ilke, Mark A. Finlayson, Paulo A. Graziano, and Anthony S. Dick. 2021. “Is There Any Incremental Benefit to Conducting Neuroimaging and Neurocognitive Assessments in the Diagnosis of ADHD in Young Children? A Machine Learning Investigation.” Developmental Cognitive Neuroscience 49 (June): 100966. https://doi.org/10.1016/j.dcn.2021.100966.

47 Palk, A. C., S. Dalvie, J. de Vries, A. R. Martin, and D. J. Stein. 2019. “Potential Use of Clinical Polygenic Risk Scores in Psychiatry – Ethical Implications and Communicating High Polygenic Risk.” Philosophy, Ethics, and Humanities in Medicine : PEHM 14 (February). https://doi.org/10.1186/s13010-019-0073-8.

48 Pedregosa, Fabian, Gaël Varoquaux, Alexandre Gramfort, Vincent Michel, Bertrand Thirion, Olivier Grisel, Mathieu Blondel, et al. 2011. “Scikit-Learn: Machine Learning in Python.” Journal of Machine Learning Research 12 (85): 2825–30.

49 Rashid, Barnaly, and Vince Calhoun. 2020. “Towards a Brain-Based Predictome of Mental Illness.” Human Brain Mapping 41 (12): 3468–3535. https://doi.org/10.1002/hbm.25013.

50 Reitan, Ralph M, and Deborah Wolfson. 1985. The Halstead-Reitan Neuropsychological Test Battery: Theory and Clinical Interpretation. Tucson, Ariz.: Neuropsychology Press.

51 Ronald, Angelica, Nora de Bode, and Tinca J. C. Polderman. 2021. “Systematic Review: How the Attention-Deficit/Hyperactivity Disorder Polygenic Risk Score Adds to Our Understanding of ADHD and Associated Traits.” Journal of the American Academy of Child & Adolescent Psychiatry 0 (0). https://doi.org/10.1016/j.jaac.2021.01.019.

52 Schachar, R., R. Tannock, M. Marriott, and G. Logan. 1995. “Deficient Inhibitory Control in Attention Deficit Hyperactivity Disorder.” Journal of Abnormal Child Psychology 23 (4): 411–37. https://doi.org/10.1007/BF01447206.

53 Simon, Alan E., Patricia N. Pastor, Cynthia A. Reuben, Larke N. Huang, and Ingrid D. Goldstrom. 2015. “Use of Mental Health Services by Children Ages Six to 11 With Emotional or Behavioral Difficulties.” Psychiatric Services (Washington, D.C.) 66 (9): 930–37. https://doi.org/10.1176/appi.ps.201400342.

54 Slobodin, Ortal, Inbal Yahav, and Itai Berger. 2020. “A Machine-Based Prediction Model of ADHD Using CPT Data.” Frontiers in Human Neuroscience 14: 383. https://doi.org/10.3389/fnhum.2020.560021.

55 Song, MinKyoung, Nathan F. Dieckmann, and Joel T. Nigg. 2019. “Addressing Discrepancies Between ADHD Prevalence and Case Identification Estimates Among U.S. Children Utilizing NSCH 2007-2012.” Journal of Attention Disorders 23 (14): 1691–1702. https://doi.org/10.1177/1087054718799930.

56 Wechsler, D. 2003. Wechsler Intelligence Scale for Children – Fourth Edition (WISC-IV). San Antonio, TX: The Psychological Corporation.

57 Wodka, Ericka L., Christopher Loftis, Stewart H. Mostofsky, Cristine Prahme, Jennifer C. Gidley Larson, Martha B. Denckla, and E. Mark Mahone. 2008. “Prediction of ADHD in Boys and Girls Using the D-KEFS.” Archives of Clinical Neuropsychology 23 (3): 283–93. https://doi.org/10.1016/j.acn.2007.12.004.

58 Youngstrom, Eric A., Tate F. Halverson, Jennifer K. Youngstrom, Oliver Lindhiem, and Robert L. Findling. 2018. “Evidence-Based Assessment from Simple Clinical Judgments to Statistical Learning: Evaluating a Range of Options Using Pediatric Bipolar Disorder as a Diagnostic Challenge.” Clinical Psychological Science: A Journal of the Association for Psychological Science 6 (2): 243–65. https://doi.org/10.1177/2167702617741845.

