## Supplemental Materials for "Prediction of ADHD diagnosis using brief, low-cost, clinical measures: a competitive model evaluation"

### **Methods**:

#### Hyperparameter Optimization:

For each of the classifiers, hyperparameters were selected within a certain range (see Table S3) based on previous work (Olson et al. 2018). An in-depth description of the parameters can be found in the *sklearn* (www.scikit-learn.org) and *bnclassify* (cran.r-project.org/package=bnclassify) documentation.

The hyperparameters were explored using a grid search and 5-fold cross-validation stratified splits. The hyperparameters with the highest mean 5-fold cross-validation accuracy for predicting best-estimate team diagnosis in the Oregon cohort were selected. These optimal hyperparameters were then used to re-train classifiers on the full training data for subsequent prediction of the Oregon test set. The same hyperparameters were used when training classifiers on the full Oregon cohort for subsequent prediction of the Michigan cohort.

#### Sensitivity Analyses:

*Classification Errors for Subthreshold and “Other” Cases:*

To better understand the low-confidence and “unstable” predictions, we examined the more granular diagnostic labels provided by the best-estimate team, which included the labels subthreshold and other. These two categories are indeed more difficult to classify overall. For instance, in the Oregon test cohort, the average class probability for subjects with a subthreshold (average stage-4 probability of 0.339) or other label (average stage-4 probability of 0.345) was significantly higher than subjects labeled as a control (average stage-4 probability of 0.048) (Supplemental Table S14). The class probabilities for subjects in these two categories also varied across the 4 stages to a greater extent than those of cases and controls, whose probabilities tended to converge across the stages (as additional data was incorporated; Figure 2). Subjects whose predicted class changed over the course of the 4 stages of the TAN classifier (i.e., class probability crossed the 0.5 threshold in subsequent stages) were more likely to be either subthreshold or other compared to subjects whose predicted class was stable (odds ratio = 4.29, Fisher’s Exact p-value = 0.000151).

When predicting KSADS diagnoses, subthreshold and other subjects were again more difficult to classify, but were more similar to ADHD cases than controls. In the Oregon test cohort, the average stage-4 class probability was 0.643 for subjects with a subthreshold KSADS label, and 0.578 for those labeled as other. Consistent with the Best-estimate team diagnosis, subjects with an unstable prediction across the TAN stages were more likely to have a subthreshold or other KSADS diagnosis (odds ratio = 4.42, Fisher’s Exact p-value = 0.0023).

*Missing Data Imputation*:

The *fancyimpute* Python package (https://github.com/iskandr/fancyimpute) was used to implement a variety of missing data imputation methods. Methods were evaluated using simulated data generated with a multivariate normal distribution with means and covariance equal to the real data. Missing values were created in the simulated data to exactly match the pattern of missingness in the real data.

The following imputation methods were implemented: k-nearest neighbor (with k=1, 3, and 7), SoftImpute (Mazumder, Hastie, and Tibshirani 2010), MatrixFactorization (https://github.com/iskandr/fancyimpute), IterativeSVD (Troyanskaya et al. 2001), and NuclearNormMinimization (Candès and Recht 2009). In Figure S2, the root mean square error (RMSE) for each method is plotted against the percent of missing data for each variable in the data set. Though NuclearNormMinimization and SoftImpute perform the best overall, KNN with k=7 was the next best performer and was chosen here due to its simplicity and ease of interpretation.

A large proportion of subjects were non-randomly missing data from the cognitive tests but this was by design—cases were advanced to cognitive testing for other studies only if they had a classifiable ADHD-non-ADHD result. Thus, we could not meaningfully disentangle missing cognitive scores from ADHD diagnostic clarity by the best-estimate team.

Nonetheless, we provide additional information here for interested readers. To determine whether missing data negatively impacted the utility of the cognitive test data for prediction, we compared classifier performance between two groups of subjects: those with data for ≥75% of the cognitive measures (N=850; N=648 in training set), and those with data for <75% of cognitive measures (N=573; N=419 in training set). Overall, mean 5-fold CV accuracy for predicting Best-estimate team diagnosis was significantly lower for subjects with missing data (80%, p=0.0018), compared to subjects with more complete cognitive data (91%). However, this is almost certainly due to the fact that a higher fraction of subjects with missing data (40%) were labeled as either subthreshold or other—those more difficult to classify—compared to subjects with more complete data (5%). To account for the confounding between missingness and diagnosis, we also examined the recall (sensitivity) for each individual diagnostic class (ADHD cases, controls, subthreshold, and other), and found no significant differences in recall between the two groups of subjects (based on missingness) for any of the four diagnostic classes (p-values 0.46 to 0.97). When predicting KSADS diagnosis, there were no significant differences between the groups for overall accuracy (p=0.95) or per-class recall (p-values 0.12 to 0.88).

*Discretization Methods*:

Sensitivity analyses were performed to determine the effect of discretizing the predictor variables. In addition to the discretization (binning) method used in all reported analyses (cut points at the mean and 1 standard deviation either side of the mean; “4bin_1st" in Table S8) we examined several other methods for choosing cut points, varying the number and size of bins, as well as

Cut points for binning were determined by two methods: (1) uniform, which results in equal distances between each cut point, and (2) percentile, which results in equal numbers of observations in each bin. In addition, clinically meaningful cut points (“Clinical” in Table S8) were also used, in which the symptom scale T-scores (ADHDRS, Conners) were categorized into low (<40), average (40-60), high-average (60-65), elevated (65-70), and very elevated (>=70) bins. The cognitive features were split along corresponding standard deviation points of -1, +1, +1.5, and +2 standard deviations resulting in 5 discretization bins. Finally, two supervised discretization methods, multi-interval discretization using the minimum description length principle (MDLP) (Fayyad and Irani 1993) and class-attribute interdependence maximization (CAIMD) (Kurgan and Cios 2004), were also implemented. For all methods, the binned variables were represented as ordinal values (e.g., 1, 2, 3, 4) when input into the classification methods.

When comparing the TAN classifier to other classifier methods, results for classifiers using both the discretized and continuous measures were evaluated (Table S8). Overall, discretization did not hinder classification performance (i.e., for the methods able to use continuous data, there was no significant difference in cross-validation accuracy when using discretized predictors vs. continuous predictors).

**Tables / Figures**:

| **Disallowed Medications** | |
|---|---|
| Abilify | Navane |
| Anafranil | Norpramine |
| Ativan | Pamelor |
| Buspar | Parnate |
| Celexa | Paxil |
| Cymbalta | Prozac |
| Depakote | Remeron |
| Effexor | Risperdal |
| Elavil/Endep | Seroquel |
| Emsam Patch | Serzone |
| Eskalith ER | Tegretol |
| Geodon | Tofranil |
| Guanfacine/Intuniv | Topamax |
| Haldol | Trazadone |
| Klonopin | Trileptal |
| Lamictil | Valium |
| Lexapro | Wellbutrin |
| Librium | Xanax |
| Lithobid | Zoloft |
| Luvox | Zyprexa |
| Nardil | |

Table S1. Disallowed medications

| Oregon-ADHD-1000 | Michigan-ADHD-1000 |
|---|---|
| **Parent-reported:** | **Parent-reported:** |
| ADHDRS: inattention | ADHDRS: inattention |
| ADHDRS: hyperactivity | ADHDRS: hyperactivity |
| Conners-3: inattention | Conners-R: cognitive problems |
| Conners-3: hyperactivity | Conners-R: hyperactivity/impulsivity |
| Conners-3: executive function | -- (not available) -- |
| Conners-3: learning problems | -- (not available) -- |
| Conners-3: aggression | -- (not available) -- |
| Conners-3: peer relations | -- (not available) -- |
| **Teacher-reported:** | **Teacher-reported:** |
| ADHDRS: inattention | ADHDRS: inattention |
| ADHDRS: hyperactivity | ADHDRS: hyperactivity |
| **Cognitive Tests:** | **Cognitive Tests:** |
| Stop-Go Task – SSRT | Stop-Go Task – SSRT |
| Stop-Go Task – GoRT | Stop-Go Task – GoRT |
| Stop-Go Task – SD GoRT | Stop-Go Task – SD GoRT |
| Spatial Span Forward | Spatial Span Forward |
| Spatial Span Backward | Spatial Span Backward |
| Digit Span Forward | Digit Span Forward |
| Digit Span Backward | Digit Span Backward |
| Stroop Task – Color | Stroop Task – Color |
| Stroop Task – Word | Stroop Task – Word |
| Stroop Task – Color-Word | Stroop Task – Color-Word |
| Trail Making Task – Condition 2 | Trail Making Task – Condition A |
| Trail Making Task – Condition 4 | Trail Making Task – Condition B |

Table S2. Predictive features used in the Oregon-ADHD-1000 and Michigan-ADHD-1000 cohorts.

| Classifier | Range of Hyperparameters | Optimal parameters: |
|---|---|---|
| TAN | score = ['loglik', 'aic', 'bic']<br>smooth = [0, 0.01, 0.1, 0.3, 0.5, 0.7, 1.0] | score = 'loglik'<br>smooth = 0.5 |
| LR | C = [0.001, 0.01, 0.1, 1, 10, 100]<br>penalty = ['l1', 'l2', 'elasticnet', 'None']<br>solver = ['liblinear', 'saga'] | C =0.1<br>penalty = 'l1'<br>solver = 'saga' |
| DT | max_depth = [1, 2, 3, 4, 5, 6, 7]<br>criterion = ['gini', 'entropy'] | criterion = 'entropy'<br>max_depth = 3 |
| RF | n_estimators = [50, 100, 200, 300, 400, 500, 800]<br>max_depth = [2, 3, 4, 5, 7]<br>max_features = [None, 'sqrt', 'auto'] | max_depth = 7<br>max_features = ‘sqrt'<br>n_estimators = 800 |
| SVM | C = [0.001, 0.01, 0.1, 1, 10, 100]<br>gamma = [0.001, 0.01, 0.1, 1, 10, 100]<br>kernel = ['linear', 'rbf', 'poly'] | C = 10<br>gamma =0.01<br>kernel = rbf' |
| GBDT | n_estimators = [50, 100, 200, 300, 400, 500, 800]<br>max_depth = [2, 3, 4, 5, 7]<br>max_features = [None, 'sqrt', 'auto'] | max_depth = 2<br>max_features = ‘sqrt'<br>n_estimators = 100 |

Table S3. Range of hyperparameters for each model and the optimized values for the discretized input method used (4 bins; cut-points at mean and 1 SD either side of the mean). A grid search was performed for each of the classifiers using 5-fold cross-validation to find optimal hyperparameters.

| Classifier | Accuracy | Specificity | Sensitivity | AUC-ROC | PPV5 | PPV50 |
|---|---|---|---|---|---|---|
| LR-unregularized | 0.851 (0.011) | 0.823 (0.027) | 0.879 (0.031) | 0.945 (0.01) | 0.21 (0.023) | 0.833 (0.019) |
| DT-Simple | 0.838 (0.012) | 0.937 (0.02) | 0.742 (0.028) | 0.909 (0.011) | 0.4 (0.087) | 0.923 (0.022) |
| TAN | 0.868 (0.014) | 0.823 (0.027) | 0.912 (0.029) | 0.948 (0.007) | 0.216 (0.023) | 0.838 (0.019) |
| LR | 0.859 (0.012) | 0.824 (0.032) | 0.891 (0.025) | 0.946 (0.009) | 0.215 (0.026) | 0.836 (0.022) |
| DT | 0.87 (0.015) | 0.876 (0.027) | 0.864 (0.035) | 0.931 (0.007) | 0.275 (0.04) | 0.875 (0.021) |
| RF | 0.884 (0.005) | 0.889 (0.028) | 0.878 (0.025) | 0.958 (0.005) | 0.304 (0.053) | 0.889 (0.023) |
| SVM | 0.878 (0.01) | 0.853 (0.024) | 0.902 (0.015) | 0.955 (0.008) | 0.248 (0.027) | 0.861 (0.018) |
| GBC | 0.89 (0.004) | 0.889 (0.015) | 0.891 (0.009) | 0.96 (0.004) | 0.301 (0.028) | 0.89 (0.013) |
| Ensemble | 0.875 (0.013) | 0.842 (0.028) | 0.908 (0.025) | 0.958 (0.005) | 0.236 (0.028) | 0.852 (0.02) |

Table S4. Mean 5-fold cross-validation performance measures for classifiers predicting Best-estimate team diagnoses in the Oregon cohort. The standard deviation of each measure across the 5 folds is given in parentheses. PPV5 = positive predictive value assuming 5% prevalence in the population; PPV50 = positive predictive value assuming 50% prevalence in the population.

| Classifier | Accuracy | Specificity | Sensitivity | AUC-ROC | PPV5 | PPV50 |
|---|---|---|---|---|---|---|
| LR-unregularized | 0.882 | 0.857 | 0.906 | 0.946 | 0.250 | 0.864 |
| DT-Simple | 0.848 | 0.937 | 0.762 | 0.904 | 0.389 | 0.924 |
| TAN | 0.874 | 0.829 | 0.917 | 0.942 | 0.220 | 0.843 |
| LR | 0.876 | 0.834 | 0.912 | 0.947 | 0.224 | 0.846 |
| DT | 0.882 | 0.891 | 0.873 | 0.924 | 0.297 | 0.889 |
| RF | 0.910 | 0.914 | 0.912 | 0.955 | 0.358 | 0.914 |
| SVM | 0.924 | 0.891 | 0.950 | 0.968 | 0.314 | 0.897 |
| GBC | 0.930 | 0.926 | 0.934 | 0.965 | 0.399 | 0.927 |
| Ensemble | 0.899 | 0.869 | 0.928 | 0.958 | 0.272 | 0.876 |

Table S5. All test-set performance measures for classifiers predicting Best-estimate team diagnosis in the Oregon cohort.

| | LR | LR-unreg | RF | DT | DT-simple | SVM | GBDT | TAN |
|---|---|---|---|---|---|---|---|---|
| LR | 1.0 (1.0) | 0.159 (0.711) | 0.023 (0.028) | 0.294 (0.500) | 0.143 (0.696) | 0.002 (0.038) | 0.004 (0.025) | 0.057 (0.033) |
| LR-unreg | | 1.0 (1.0) | 0.011 (0.006) | 0.086 (0.355) | 0.245 (0.858) | 0.008 (0.011) | **0.002 (0.004)** | 0.027 (0.012) |
| RF | | | 1.0 (1.0) | 0.07 (0.109) | 0.001 (0.016) | 0.245 (0.120) | 0.108 (0.487) | 0.221 (0.068) |
| DT | | | | 1.0 (1.0) | 0.005 (0.230) | 0.397 (0.512) | 0.054 (0.091) | 0.388 (0.260) |
| DT-simple | | | | | 1.0 (1.0) | 0.015 (0.103) | **0.002 (0.006)** | 0.014 (0.035) |
| SVM | | | | | | 1.0 (1.0) | 0.031 (0.212) | 0.758 (0.259) |
| GBDT | | | | | | | 1.0 (1.0) | 0.056 (0.751) |
| TAN | | | | | | | | 1.0 (1.0) |

Table S6. P-values from paired T-tests comparing classifier accuracy for predicting best-estimate team diagnosis in the Oregon cohort. P-values for tests comparing both 5-fold cross-validation accuracies and 5x2-fold cross-validation accuracies (in parentheses) are reported. Only gradient boosted decision trees (GBDT) showed a consistent significant improvement ($p<0.007$) over unregularized logistic regression and the 2-level decision tree for both cross-validation methods (bolded values).

| Classifier | 5-fold CV (SD) | Test Set |
|---|---|---|
| DT-simple | 0.866 (0.017) | 0.868 |
| LR-unregularized | 0.857 (0.023) | 0.879 |
| LR | 0.866 (0.028) | 0.890 |
| DT | 0.866 (0.017) | 0.868 |
| RF | 0.891 (0.018 | 0.896 |
| SVM | 0.889 (0.013) | 0.885 |
| GBDT | 0.892 (0.017) | 0.904 |
| Ensemble | 0.897 (0.006) | 0.904 |

Table S7. Mean 5-fold cross validation accuracies for classifiers predicting best-estimate team diagnosis in the Oregon cohort. All classifiers used the full parent, teacher, and EF feature set with **continuous** values.

| Classifier | 4 bins percentile | 4 bins uniform | 6 bins percentile | 8 bins percentile | Continuous* | 4bin_1st | MDLP | CAIMD | Clinical |
|---|---|---|---|---|---|---|---|---|---|
| DT-simple | 0.828 | 0.828 | 0.83 | 0.856 | 0.866 | 0.84 | 0.849 | 0.835 | 0.845 |
| TAN | 0.87 | 0.868 | 0.867 | 0.874 | NA | 0.868 | 0.886 | 0.86 | 0.841 |
| LR | 0.881 | 0.866 | 0.878 | 0.883 | 0.866 | 0.858 | 0.882 | 0.869 | 0.845 |
| DT | 0.872 | 0.871 | 0.863 | 0.872 | 0.866 | 0.87 | 0.87 | 0.873 | 0.855 |
| RF | 0.902 | 0.883 | 0.885 | 0.888 | 0.891 | 0.884 | 0.896 | 0.886 | 0.861 |
| SVM | 0.892 | 0.879 | 0.891 | 0.903 | 0.889 | 0.877 | 0.891 | 0.876 | 0.864 |
| GBDT | 0.904 | 0.906 | 0.897 | 0.899 | 0.892 | 0.89 | 0.896 | 0.886 | 0.861 |

Table S8. Accuracy measures for classifiers across a variety of methods (see descriptions above) for discretization of predictors. Hyperparameters were optimized for each input method and classifier. *There were no statistically significant differences in accuracy between the models using continuous vs. discretized features. The “4bin_1st" column represents the method use in all analyses reported in the paper (cut points at the mean and 1 standard deviation either side of the mean.

| | | Best-estimate Team Diagnosis | | | | |
|---|---|---|---|---|---|---|
| | | Control | Subthreshold | ADHD | Not-clean control | Total |
| KSADS Diagnosis | Control | 402 | 57 | 28 | 83 | 570 |
| | Subthreshold | 5 | 11 | 76 | 11 | 103 |
| | ADHD | 2 | 7 | 571 | 56 | 636 |
| | Not-clean control | 12 | 31 | 49 | 21 | 113 |
| | Total | 421 | 106 | 724 | 171 | |

Table S9. Agreement between KSADS and Best-estimate team labels for the Oregon-ADHD-1000 (N=1422 have both labels).

| **Classifier** | **5-fold CV (SD)** | **Test Set** | **Test Set, High-confidence** |
|---|---|---|---|
| LR-unregularized | 0.830 (0.015) | 0.829 | 0.947 (N=170, 48%) |
| DT-simple | 0.843 (0.024) | 0.840 | 0.951 (N=145, 41%)* |
| TAN-stage4 | 0.848 (0.014) | 0.820 | 0.860 (N=328, 92%) |
| TAN-earliest | 0.849 (0.015) | 0.820 | 0.860 (N=329, 92%) |
| LR | 0.828 (0.011) | 0.834 | 0.976 (N=126, 35%) |
| DT | 0.832 (0.025) | 0.815 | 0.952 (N=147, 41%)* |
| RF | 0.844 (0.017) | 0.831 | 0.963 (N=135, 38%) |
| SVM | 0.838 (0.017) | 0.837 | 0.954 (N=130, 37%) |
| GBDT | 0.842 (0.014) | 0.848 | 0.947 (N=171, 48%) |
| Ensemble | 0.851 (0.011) | 0.829 | 0.971 (N=138, 39%) |

Table S10. Mean 5-fold cross-validation accuracies for classifiers predicting KSADS diagnoses in the Oregon cohort. The test set is the Oregon hold-out sample (N=356). Also shown (last column) are the accuracies for the subset of the test set with high-confidence predictions (predicted class probability >0.9); the number of high-confidence predictions and the corresponding percentage of the test set are given in parentheses. The classifiers included all discretized parent, teacher, and cognitive predictive features. *These models were able to make high-confidence predictions of controls only (i.e., high-confidence sensitivity=0).

| Classifier | Accuracy | Specificity | Sensitivity | AUC-ROC | PPV5 | PPV50 |
|---|---|---|---|---|---|---|
| LR-unregularized | 0.83 (0.015) | 0.795 (0.031) | 0.874 (0.037) | 0.908 (0.013) | 0.186 (0.02) | 0.811 (0.02) |
| DT-Simple | 0.843 (0.024) | 0.81 (0.023) | 0.884 (0.033) | 0.878 (0.015) | 0.199 (0.027) | 0.823 (0.022) |
| TAN | 0.848 (0.012) | 0.805 (0.012) | 0.901 (0.018) | 0.911 (0.017) | 0.196 (0.011) | 0.822 (0.011) |
| LR | 0.828 (0.011) | 0.78 (0.029) | 0.889 (0.034) | 0.914 (0.013) | 0.177 (0.015) | 0.802 (0.016) |
| DT | 0.832 (0.025) | 0.8 (0.036) | 0.872 (0.047) | 0.886 (0.009) | 0.19 (0.028) | 0.814 (0.026) |
| RF | 0.844 (0.017) | 0.808 (0.024) | 0.889 (0.026) | 0.917 (0.011) | 0.198 (0.02) | 0.823 (0.019) |
| SVM | 0.838 (0.012) | 0.781 (0.033) | 0.908 (0.017) | 0.905 (0.01) | 0.182 (0.019) | 0.807 (0.021) |
| GBDT | 0.842 (0.014) | 0.81 (0.022) | 0.882 (0.024) | 0.915 (0.011) | 0.198 (0.017) | 0.823 (0.016) |
| Ensemble | 0.851 (0.011) | 0.807 (0.015) | 0.905 (0.015) | 0.917 (0.01) | 0.199 (0.013) | 0.824 (0.011) |

Table S11. All mean 5-fold cross-validation performance measures for classifiers predicting KSADS diagnosis in the Oregon cohort.

| Classifier | Accuracy | Specificity | Sensitivity | AUC-ROC | PPV5 | PPV50 |
|---|---|---|---|---|---|---|
| LR-unregularized | 0.829 | 0.796 | 0.869 | 0.904 | 0.183 | 0.810 |
| DT-Simple | 0.840 | 0.791 | 0.900 | 0.864 | 0.185 | 0.811 |
| TAN-stage4 | 0.837 | 0.796 | 0.888 | 0.905 | 0.186 | 0.813 |
| LR | 0.834 | 0.791 | 0.888 | 0.902 | 0.183 | 0.809 |
| DT | 0.815 | 0.811 | 0.819 | 0.879 | 0.186 | 0.813 |
| RF | 0.831 | 0.796 | 0.875 | 0.901 | 0.184 | 0.811 |
| SVM | 0.834 | 0.781 | 0.900 | 0.903 | 0.178 | 0.804 |
| GBDT | 0.848 | 0.827 | 0.875 | 0.905 | 0.210 | 0.835 |
| Ensemble | 0.837 | 0.796 | 0.888 | 0.905 | 0.186 | 0.813 |

Table S12. All test-set performance measures for models predicting KSADS diagnosis in the Oregon cohort.

| | LR | LR-unreg | RF | DT | DT-simple | SVM | GBDT | TAN |
|---|---|---|---|---|---|---|---|---|
| LR | 1.0 (1.0) | 0.647 (0.091) | 0.091 (0.055) | 0.822 (0.071) | 0.303 (0.185) | 0.248 (0.256) | 0.137 (0.227) | 0.018 (0.065) |
| LR-unreg | | 1.0 (1.0) | 0.153 (0.020) | 0.916 (0.031) | 0.423 (0.049) | 0.294 (0.112) | 0.173 (0.064) | 0.037 (0.008) |
| RF | | | 1.0 (1.0) | 0.321 (0.741) | 0.925 (0.224) | 0.469 (0.192) | 0.690 (0.001) | 0.135 (0.134) |
| DT | | | | 1.0 (1.0) | 0.242 (0.216) | 0.694 (0.17) | 0.340 (0.085) | 0.148 (0.192) |
| DT-simple | | | | | 1.0 (1.0) | 0.638 (0.504) | 0.932 (0.573) | 0.505 (0.457) |
| SVM | | | | | | 1.0 (1.0) | 0.509 (0.600) | 0.147 (0.700) |
| GBDT | | | | | | | 1.0 (1.0) | 0.037 (0.708) |
| TAN | | | | | | | | 1.0 (1.0) |

Table S13. P-values from paired T-tests comparing classifier accuracy for predicting KSADS diagnosis in the Oregon cohort. P-values for tests comparing both 5-fold cross-validation accuracies and 5x2-fold cross-validation accuracies (in parentheses) are reported. No classifier showed consistent significant improvement over any other.

| | Stage 1 | | Stage 2 | | Stage 3 | | Stage 4 | |
|---|---|---|---|---|---|---|---|---|
| | CV | Test | CV | Test | CV | Test | CV | Test |
| ADHD cases | 0.759 | 0.739 | 0.870 | 0.839 | 0.901 | 0.885 | 0.909 | 0.906 |
| Controls | 0.135 | 0.105 | 0.0902 | 0.0698 | 0.0566 | 0.0561 | 0.0450 | 0.0478 |
| Subthreshold | 0.444 | 0.440 | 0.404 | 0.452 | 0.399 | 0.414 | 0.353 | 0.339 |
| Other | 0.433 | 0.497 | 0.445 | 0.502 | 0.419 | 0.438 | 0.374 | 0.345 |

Table S14. Average class probabilities at each stage of the TAN classifier. All probabilities shown are the probability of an ADHD diagnosis.

| | | Best-estimate Team Diagnosis | | | | |
|---|---|---|---|---|---|---|
| | | Control | Subthreshold | ADHD | Other | Total |
| KSADS Diagnosis | Control | 413 | 34 | 6 | NA | 453 |
| | Subthreshold | 8 | 16 | 26 | NA | 50 |
| | ADHD | 31 | 48 | 414 | NA | 493 |
| | Other | 19 | 28 | 12 | NA | 59 |
| | Total | 471 | 126 | 458 | NA | |

Table S15. Agreement between KSADS and Best-estimate team labels for the Michigan-ADHD-1000 (N=1055 have both labels).

| Classifier | Training Set (Oregon; N=1422) | Test Set (Michigan; N=1055) | Test Set (Michigan; High-confidence) |
|---|---|---|---|
| LR-unregularized | 0.838 | 0.741 | 0.864 (N=544, 52%) |
| DT-simple | 0.846 | 0.755 | 0.742 (N=675, 64%)* |
| TAN-stage4 | 0.878 | 0.789 | 0.821 (N=950, 90%) |
| LR | 0.835 | 0.750 | 0.883 (N=511, 48%)* |
| DT | 0.846 | 0.755 | 0.888 (N=482, 46%)* |
| RF | 0.899 | 0.761 | 0.918 (N=449, 43%) |
| SVM | 0.844 | 0.758 | 0.861 (N=532, 50%)* |
| GBDT | 0.858 | 0.730 | 0.849 (N=569, 54%) |
| Ensemble | 0.860 | 0.767 | 0.872 (N=531, 50%)* |

Table S16. Accuracy of the classifiers predicting KSADS diagnoses in the Michigan cohort. The classifiers were trained on the Oregon cohort data, using all discretized parent, teacher and cognitive predictive features. *These models were able to make high-confidence predictions of controls only (i.e., high-confidence sensitivity = 0).

| Classifier | Accuracy | Specificity | Sensitivity | PPV5 | PPV50 | AUC-ROC |
|---|---|---|---|---|---|---|
| LR-unregularized | 0.779 | 0.890 | 0.633 | 0.881 | 0.232 | 0.852 |
| DT-Simple | 0.754 | 0.912 | 0.548 | 0.850 | 0.246 | 0.861 |
| TAN-stage4 | 0.792 | 0.883 | 0.672 | 0.874 | 0.232 | 0.852 |
| LR | 0.785 | 0.888 | 0.651 | 0.884 | 0.234 | 0.853 |
| DT | 0.770 | 0.861 | 0.651 | 0.857 | 0.198 | 0.824 |
| RF | 0.807 | 0.893 | 0.694 | 0.889 | 0.255 | 0.867 |
| SVM | 0.805 | 0.885 | 0.701 | 0.883 | 0.243 | 0.859 |
| GBC | 0.822 | 0.890 | 0.734 | 0.892 | 0.259 | 0.869 |
| Ensemble | 0.807 | 0.893 | 0.694 | 0.890 | 0.255 | 0.867 |

Table S17. All test-set performance measures for classifiers predicting best-estimate team diagnosis in the Michigan cohort.

| Classifier | Accuracy | Specificity | Sensitivity | PPV5 | PPV50 | AUC-ROC |
|---|---|---|---|---|---|---|
| LR-unregularized | 0.741 | 0.938 | 0.517 | 0.903 | 0.304 | 0.893 |
| DT-Simple | 0.755 | 0.934 | 0.552 | 0.778 | 0.306 | 0.893 |
| TAN-stage4 | 0.789 | 0.899 | 0.663 | 0.881 | 0.256 | 0.867 |
| LR | 0.750 | 0.936 | 0.538 | 0.905 | 0.306 | 0.894 |
| DT | 0.766 | 0.925 | 0.584 | 0.866 | 0.291 | 0.887 |
| RF | 0.761 | 0.932 | 0.566 | 0.901 | 0.306 | 0.893 |
| SVM | 0.758 | 0.922 | 0.572 | 0.901 | 0.278 | 0.880 |
| GBC | 0.730 | 0.950 | 0.479 | 0.906 | 0.336 | 0.906 |
| Ensemble | 0.767 | 0.927 | 0.584 | 0.907 | 0.296 | 0.889 |

Table S18. All test-set performance measures for classifiers predicting KSADS diagnosis in the Michigan cohort.

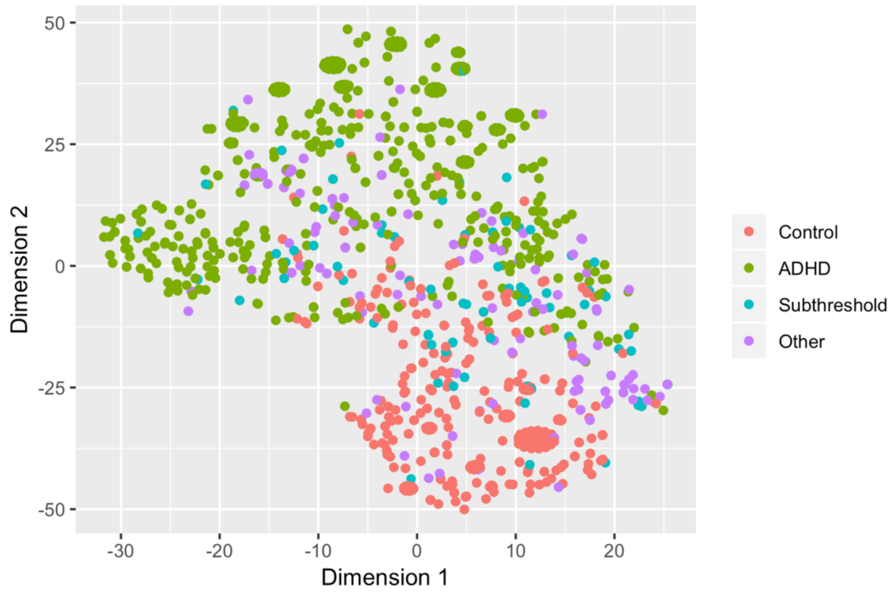


Figure S1. A 2-dimensional t-distributed stochastic neighbor embedding (t-SNE) visualization of all participants in the Oregon-ADHD-1000 cohort training set, showing fairly good separation between ADHD cases and controls but significant heterogeneity among the other categories. Dimensionality reduction was performed on the 10 parent and teacher ratings scales (cognitive measures were not included here). Parameters used for the t-SNE operation were: perplexity=30, distance=Euclidean.

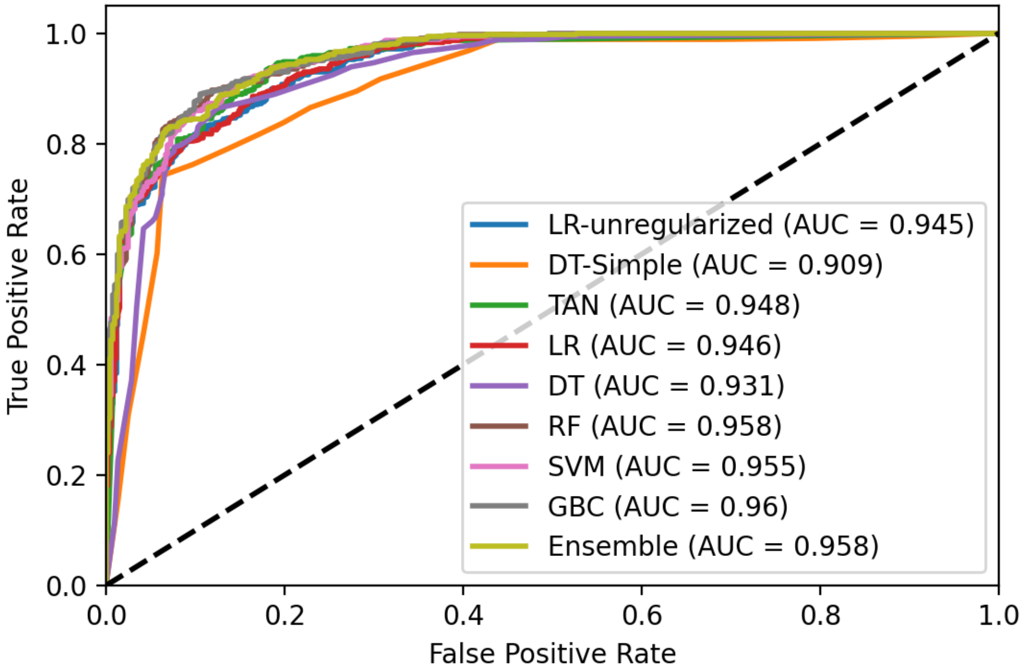


Figure S2. Receiver operating characteristic (ROC) curves showing the mean 5-fold cross-validation performance for all classifiers predicting best-estimate team diagnosis.

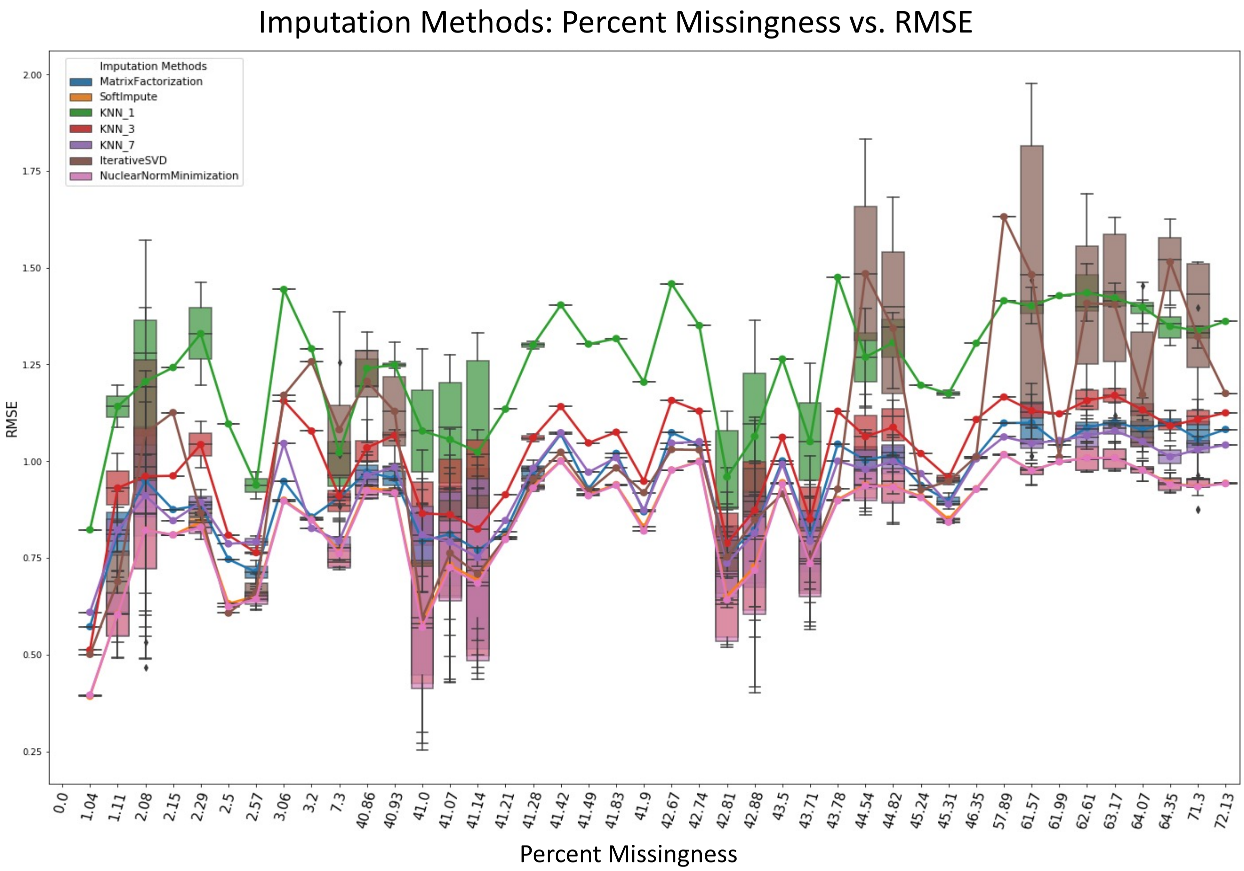


Figure S3. Performance of missing data imputation methods. The root mean square error of seven imputation methods is shown across predictive features with a wide range of missingness.

References for the Supplement:

Candès, Emmanuel J., and Benjamin Recht. 2009. "Exact Matrix Completion via Convex Optimization." *Foundations of Computational Mathematics* 9 (6): 717. https://doi.org/10.1007/s10208-009-9045-5.

Fayyad, U., and K. Irani. 1993. "Multi-Interval Discretization of Continuous-Valued Attributes for Classification Learning," September. https://trs.jpl.nasa.gov/handle/2014/35171.

Kurgan, L.A., and K.J. Cios. 2004. "CAIM Discretization Algorithm." *IEEE Transactions on Knowledge and Data Engineering* 16 (2): 145–53. https://doi.org/10.1109/TKDE.2004.1269594.

Mazumder, Rahul, Trevor Hastie, and Robert Tibshirani. 2010. "Spectral Regularization Algorithms for Learning Large Incomplete Matrices." *Journal of Machine Learning Research: JMLR* 11 (March): 2287–2322.

Olson, Randal S., William La Cava, Zairah Mustahsan, Akshay Varik, and Jason H. Moore. 2018. "Data-Driven Advice for Applying Machine Learning to Bioinformatics Problems." *Pacific Symposium on Biocomputing. Pacific Symposium on Biocomputing* 23: 192–203.

Troyanskaya, O., M. Cantor, G. Sherlock, P. Brown, T. Hastie, R. Tibshirani, D. Botstein, and R. B. Altman. 2001. "Missing Value Estimation Methods for DNA Microarrays." *Bioinformatics (Oxford, England)* 17 (6): 520–25. https://doi.org/10.1093/bioinformatics/17.6.520.
